# Advances in methods for characterizing dietary patterns: A scoping review

**DOI:** 10.1101/2024.06.20.24309251

**Authors:** Joy M. Hutchinson, Amanda Raffoul, Alexandra Pepetone, Lesley Andrade, Tabitha E. Williams, Sarah A. McNaughton, Rebecca M. Leech, Jill Reedy, Marissa M. Shams-White, Jennifer E. Vena, Kevin W. Dodd, Lisa M. Bodnar, Benoît Lamarche, Michael P. Wallace, Megan Deitchler, Sanaa Hussain, Sharon I. Kirkpatrick

**Affiliations:** School of Public Health Sciences, University of Waterloo, Waterloo, ON, Canada; Department of Nutritional Sciences, University of Toronto, Toronto, ON, Canada; Health and Well-Being Centre for Research Innovation, School of Human Movement and Nutrition Sciences, University of Queensland, St. Lucia, QLD, Australia; Institute for Physical Activity and Nutrition, School of Exercise and Nutrition Sciences, Deakin University, Victoria, Geelong, Australia; National Cancer Institute, National Institutes of Health, Bethesda, MD, USA; Population Science Department, American Cancer Society, Washington DC, USA; Division of Cancer Control and Population Sciences, National Cancer Institute, Bethesda, MD, USA; Alberta’s Tomorrow Project, Alberta Health Services, Edmonton, AB, Canada; Division of Cancer Prevention, National Cancer Institute, Bethesda, MD, USA; School of Public Health, University of Pittsburgh, Pittsburgh, PA, USA; Centre Nutrition, santé et société (NUTRISS), Institut sur la nutrition et les aliments fonctionnels (INAF), Université Laval, Québec City, QC, Canada; Department of Statistics and Actuarial Science, University of Waterloo, Waterloo, ON, Canada; Intake – Center for Dietary Assessment, FHI Solutions, Washington, DC, USA

## Abstract

There is a growing focus on better understanding the complexity of dietary patterns and how they relate to health and other factors. Approaches that have not traditionally been applied to characterize dietary patterns, such as machine learning algorithms and latent class analysis methods, may offer opportunities to measure and characterize dietary patterns in greater depth than previously considered. However, there has not been a formal examination of how this wide range of approaches has been applied to characterize dietary patterns. This scoping review synthesized literature from 2005-2022 applying methods not traditionally used to characterize dietary patterns, referred to as novel methods. MEDLINE, CINAHL, and Scopus were searched using keywords including machine learning, latent class analysis, and least absolute shrinkage and selection operator (LASSO). Of 5274 records identified, 24 met the inclusion criteria. Twelve of 24 articles were published since 2020. Studies were conducted across 17 countries. Nine studies used approaches that have applications in machine learning to identify dietary patterns. Fourteen studies assessed associations between dietary patterns that were characterized using novel methods and health outcomes, including cancer, cardiovascular disease, and asthma. There was wide variation in the methods applied to characterize dietary patterns and in how these methods were described. The extension of reporting guidelines and quality appraisal tools relevant to nutrition research to consider specific features of novel methods may facilitate complete and consistent reporting and enable evidence synthesis to inform policies and programs aimed at supporting healthy dietary patterns.

## Introduction

Dietary intake is among the top risk factors for chronic diseases.^1,2^ Research examining dietary intake has historically focused on single foods, nutrients, or other dietary constituents.^3^ As the focus of public health nutrition shifted from the prevention of deficiency to include a focus on the prevention of chronic diseases, research likewise shifted towards the examination of dietary patterns, aiming to capture how foods and beverages are consumed in real life.^3–5^ Humans typically do not consume foods or nutrients on their own, but in the context of a broader dietary pattern.^3,4^ Accordingly, food-based dietary guidelines are now typically focused on patterns of intake rather than single dietary components.^6^ It is likely the synergistic and antagonistic relationships among the multiple foods, beverages, and other dietary components that humans consume that influence health rather than individual components.^4^ In addition to this multidimensionality, dietary patterns are dynamic, changing from meal to meal, day to day and across the life course.^4,7^ Further, dietary patterns are shaped by culture, social position, and other contextual factors.^8,9^ However, incorporating the domains of multidimensionality, dynamism, and contextual factors into dietary patterns analysis is a difficult task.

Traditional approaches to identify dietary patterns, including ‘a priori’ and ‘a posteriori’ approaches, are useful for understanding overall dietary patterns or diet quality of populations and population subgroups.^10^ For example, ‘a priori’ methods like the Healthy Eating Index-2020 or the Healthy Eating Food Index-2019 are generally investigator driven,^11,12^ consider multiple components as inputs, such as fruits and vegetables and whole grains, but typically compress the multidimensional construct of total dietary patterns, condensing inputs to a single unidimensional score reflecting overall diet quality.^13,14^ ‘A posteriori’ approaches are data-driven and have also been widely used to identify dietary patterns. Commonly applied data-driven approaches include clustering methods (e.g., k-means, Ward’s method), principal component analysis, and factor analysis, providing opportunities to identify dietary patterns through statistical modelling or clustering algorithms rather than relying on researcher hypotheses.^15^ These approaches compress dietary components to key food groupings typically expressed as single scores.^10,16^ By reducing the dimensionality of dietary patterns, these methods are limited in their ability to explain the wide variation in dietary intakes.^4^ Methods employed to traditionally characterize dietary patterns using ‘a priori’ and ‘a posteriori’ approaches thus address multidimensionality to some extent, but do not allow for explorations of dietary patterns in their totality because they miss potential synergistic or antagonistic associations among dietary components.^4,14,17^

Novel methods that have not traditionally been used to identify dietary patterns, such as probabilistic graphical modelling, latent class analysis, and machine learning algorithms (e.g., random forest, neural networks), may capture complexities like dietary synergy. There is no clear delineation between traditional and novel methods, and specifically defining what is novel is challenging given it naturally implies an evolution of methods. Nonetheless, there is a growing interest among nutrition researchers in the application of methods that have not typically been used to capture dietary complexity, with these methods often centered in machine learning.^18^ To date, there have been perspectives and narrative reviews on the application of machine learning in nutrition,^19–21^ and a recent systematic review of studies that applied machine learning approaches to assess food consumption.^22^ However, there has not been an assessment of studies applying novel methods to characterize dietary patterns. Given the rapid adoption of these methods within the field of health,^23–26^ it is increasingly important for researchers to have a basic understanding of available methods and how they are being applied in the field. This will facilitate the synthesis of evidence from a range of methodological inputs to inform food-based dietary guidelines and other policies and programs that promote health. The objective of this scoping review was therefore to describe the use of novel methods not traditionally used to characterize dietary patterns in the published literature.

## Methods

The review was conducted in accordance with the JBI Manual for Evidence Synthesis,^27^ which was developed using the Arksey and O’Malley framework.^28^ Reporting follows the Preferred Reporting Items for Systematic Reviews and Meta-Analyses extension for Scoping Reviews (PRISMA-ScR).^29^

### Defining novel methods

The novel methods considered were based on a preliminary search of the literature and the expertise of the research team and included systems methods (e.g., agent-based modelling, system dynamics), least absolute shrinkage and selection operator (LASSO), machine learning algorithms, copulas, and data-driven statistical modelling approaches (e.g., treelet transformations, principal balances and coordinates). Novel methods could also include those that have been used previously in nutrition research if applied in new ways to characterize dietary patterns (e.g., linear programming used to model a modified dietary pattern rather than to test scenarios). Methods that were not considered to be novel were those that have been applied to assess dietary patterns in numerous studies and have been considered by prior reviews and commentaries,^2,10,30^ including regression, ‘a priori’ approaches such as investigator-driven indices, and routinely used data-driven approaches, including factor analysis and cluster analysis.^10,31^

### Identifying relevant studies

Articles were eligible for inclusion if they were: a primary research article; focused on dietary intake as an exposure or outcome, including examination of dietary patterns (i.e., multiple dietary components in combination rather than single nutrients, foods, or other dietary components); used at least one or more novel methods to characterize dietary patterns as described above; were published in English; and focused on humans. Ineligible studies included those focused on individual foods or human milk rather than dietary patterns, and commentaries and reviews.

Searches of three research databases, MEDLINE (via PubMed), the Cumulative Index to Nursing and Allied Health Literature (CINAHL), and Scopus, were conducted in March 2022. These health-focused, specialized, and multidisciplinary databases were selected based on consultation with a research librarian (JS) to ensure a range of possibly relevant study types were included. The search strategies were developed in consultation with the research librarian using keywords and subject headings to capture diet-related constructs (e.g., dietary intake, patterns, recommendations, feeding behaviour, food habits) and novel methods to characterize dietary patterns (e.g., machine learning, network science, system dynamics model). No date limits were applied to the searches, and articles were included until the end of the search in March 2022. The search strategies for MEDLINE, CINAHL, and Scopus are available in **Supplemental File 1**.

### Study selection

Two independent reviewers (two of AP, AR, SH, SIK) screened each record at the title and abstract and full-text screening stages using Covidence,^32^ with one consistent reviewer (AP) participating throughout the entire process. At the title and abstract screening stage, an initial pilot screening (25 records) generated 100% agreement (AR and AP) and 92% agreement (AP and SH). A second pilot screening (100 records) generated 91% agreement (AR and AP) and 93% agreement (AP and SH). When applicable, discrepancies were discussed by reviewers and if needed deferred to a third reviewer (SIK) for decision. Following pilot screening, the reviewers independently reviewed the remaining articles (96% agreement, Kappa = 0.83).

The reviewers were intentionally liberal during the title and abstract screening stage because of the breadth of possible novel methods. This required iteratively revisiting the inclusion criteria. For example, reduced rank regression was initially considered to be novel but was found to be prevalent in the literature based on title and abstract screening and was excluded during full-text review. Further, articles that used ‘a posteriori’ methods to identify dietary intake but did not specify the exact method in the title or abstract were included for full-text review.

Pilot screening of full-text reviews (50 records) generated 82% agreement (AR and AP) and 96% agreement (AP and SIK); after discrepancies were discussed, two reviewers independently screened the remaining full-text articles (93% agreement, Kappa = 0.60). The high agreement between reviewers but relatively low Cohen’s Kappa is described as Cohen’s paradox, with a larger number of studies excluded than included.^33–35^

### Data extraction

Data extraction was completed by TEW and JMH using a pre-specified Excel template, with all extracted data subsequently verified by LA. Data extraction fields (**Supplemental File 2**) included information pertaining to authorship, study title, journal, year of publication, funding source, contextual details (e.g., study location), sample size, and participant characteristics (e.g., age). Details relating to study methods (e.g., analysis input variables, measurement of dietary intake, analytic approaches) and results (e.g., findings related to dietary patterns and if applicable, health risk and outcomes) were also extracted.

## Results

### Summary of search

A total of 5274 unique articles were identified after removing duplicates. Of these, 436 were identified as potentially relevant based on the title and abstract review and underwent full-text screening (**Figure 1**). Studies excluded during full-text screening included those that did not include methods defined as novel, those that did not focus on dietary patterns, commentaries, narrative reviews, systematic reviews, studies that were not published in English, studies that were not conducted with humans, and theses/dissertations. A final pool of 24 articles describing 24 unique studies met the inclusion criteria.

**Figure 1.**
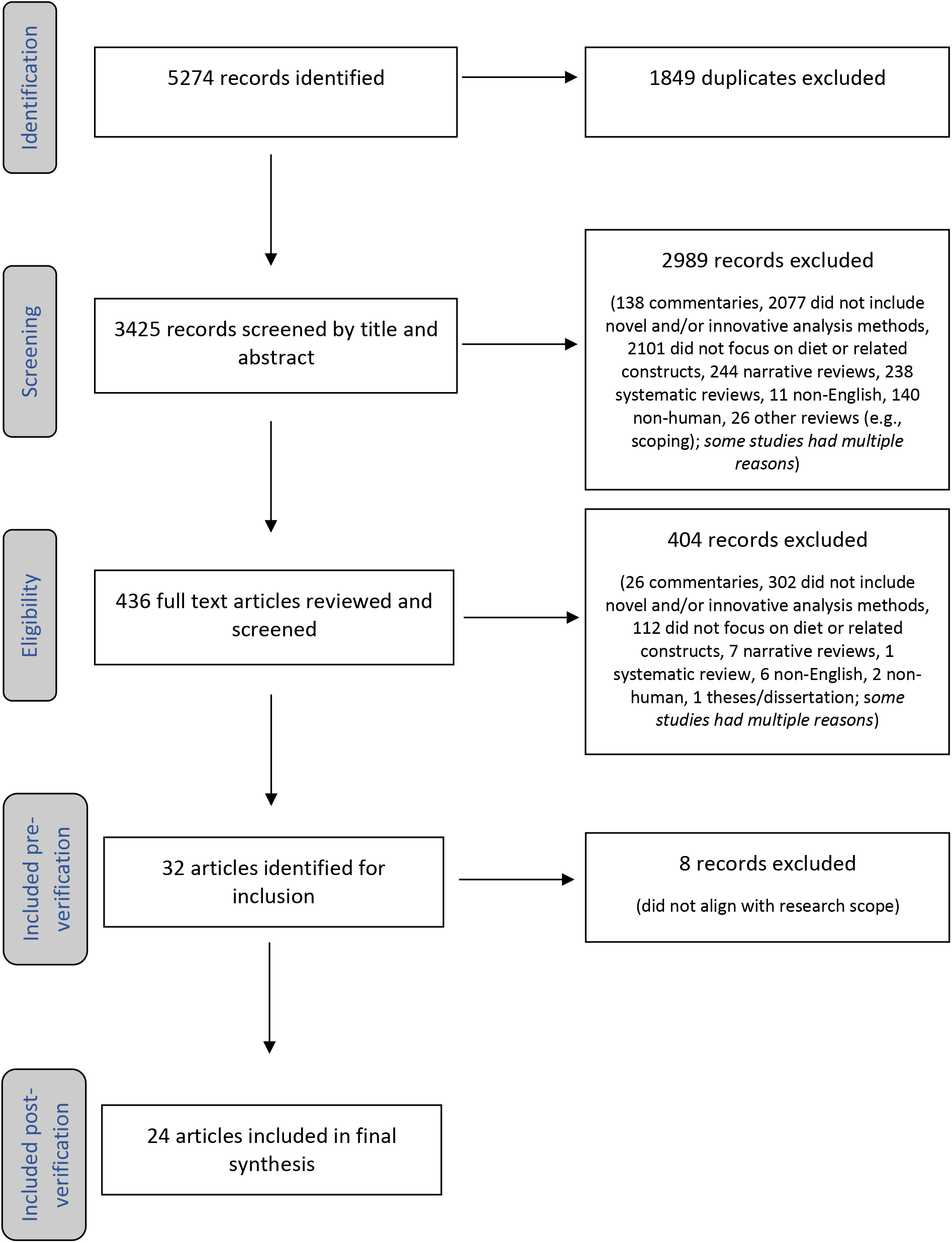
PRISMA diagram illustrating the screening process for a scoping review exploring innovative methods for the analysis of dietary intake data and characterization of dietary patterns.

### Characteristics of included studies

Across the 24 included studies, data from 17 countries were represented (**Table 1**). Half of the studies were published between 2005 and 2019,^36–47^ and the remaining 12 were published between 2020 and March 2022.^48–59^ Three studies used data from subsets of the European Prospective Investigation into Cancer and Nutrition,^36,45,46^ two studies used waves of data from the National Health and Nutrition Examination Survey,^54,58^ and two studies used data from the ELSA-Brasil cohort study (**Table 2**).^42,57^ Sample sizes ranged from 250 to over 73,000 participants. Nineteen studies were conducted using data from cohort or cross-sectional studies and five studies applied a case-control design.

**Table 1:**
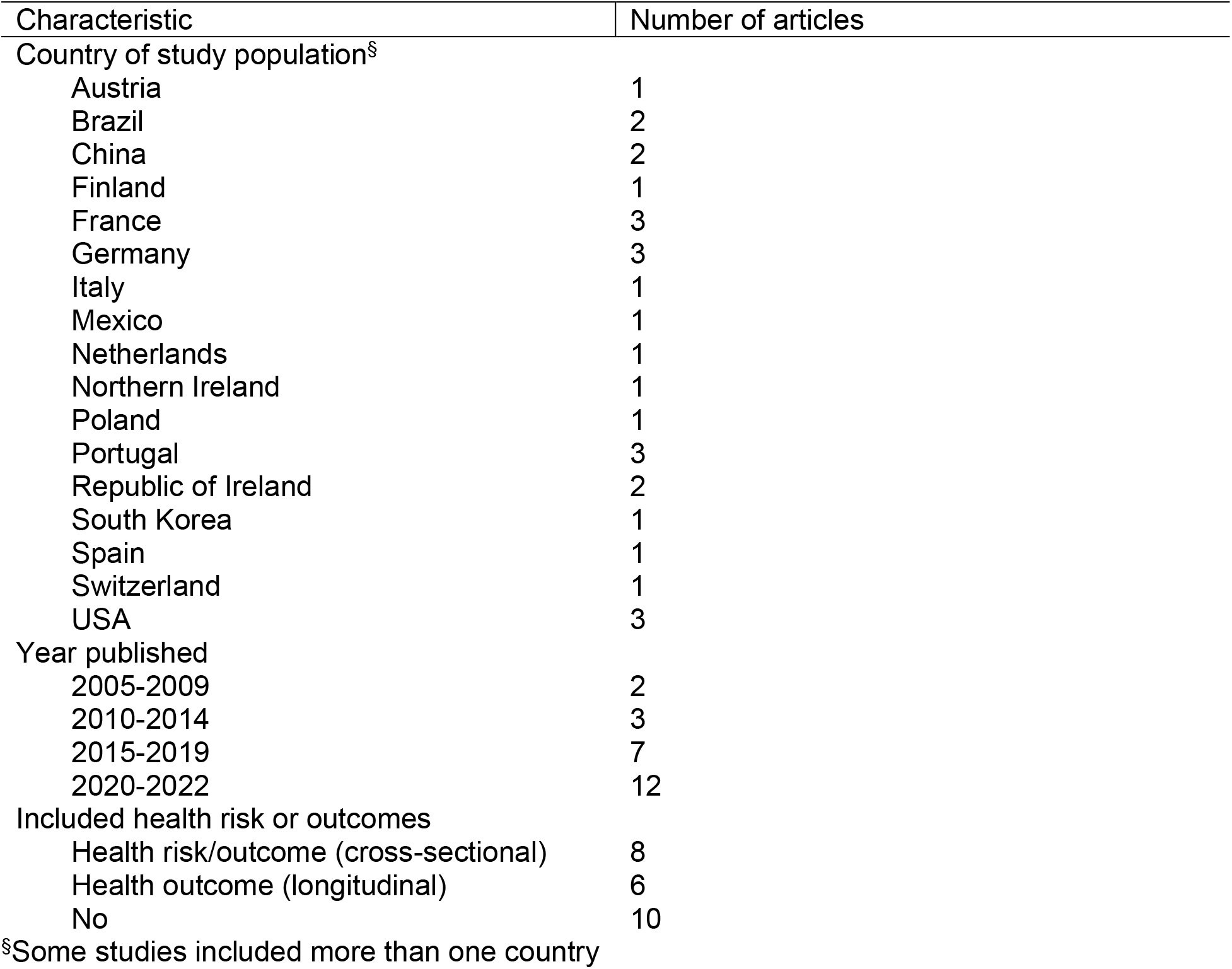
Study characteristics across included studies applying novel methods to characterize dietary patterns.

**Table 2:**
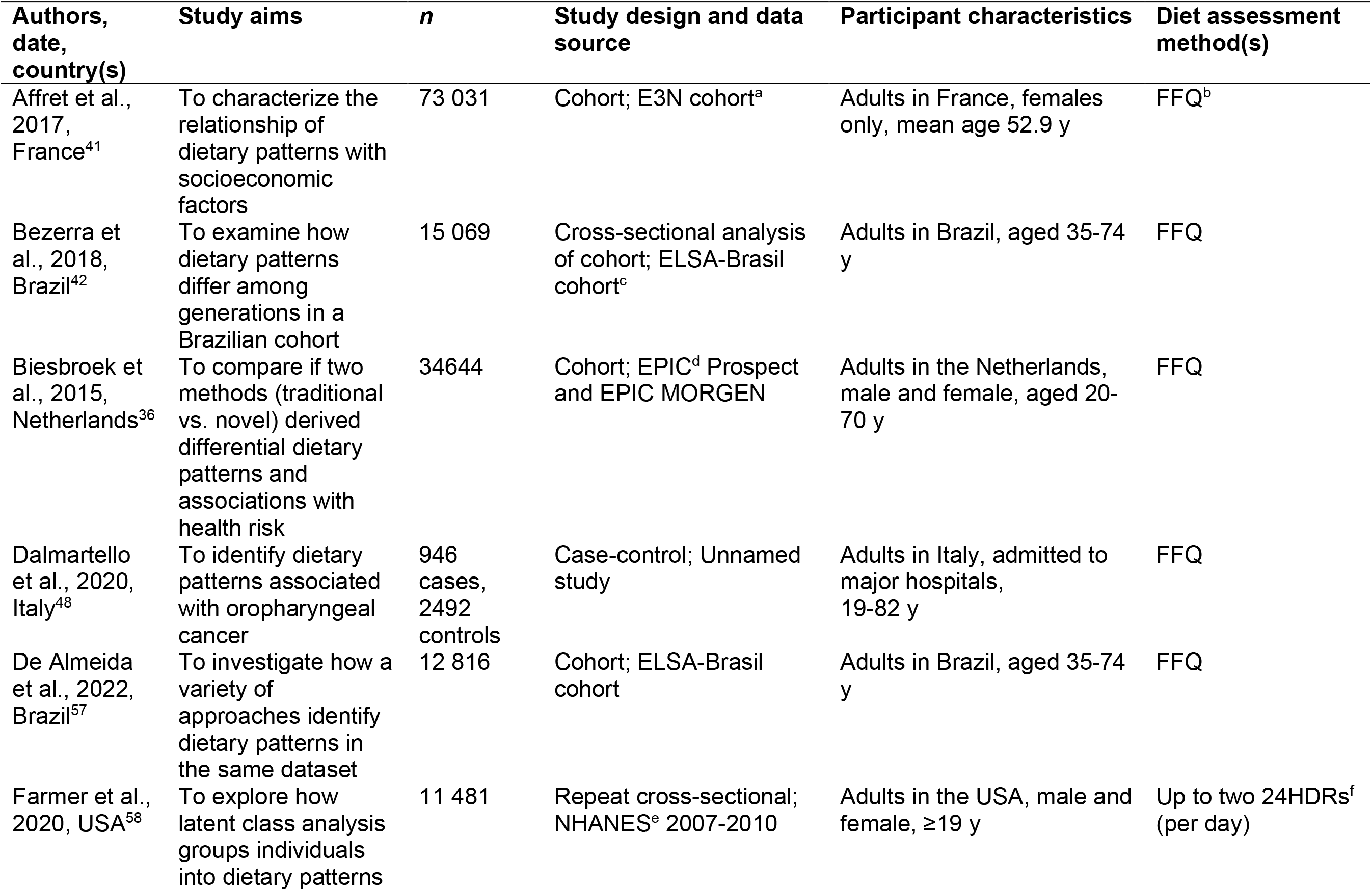
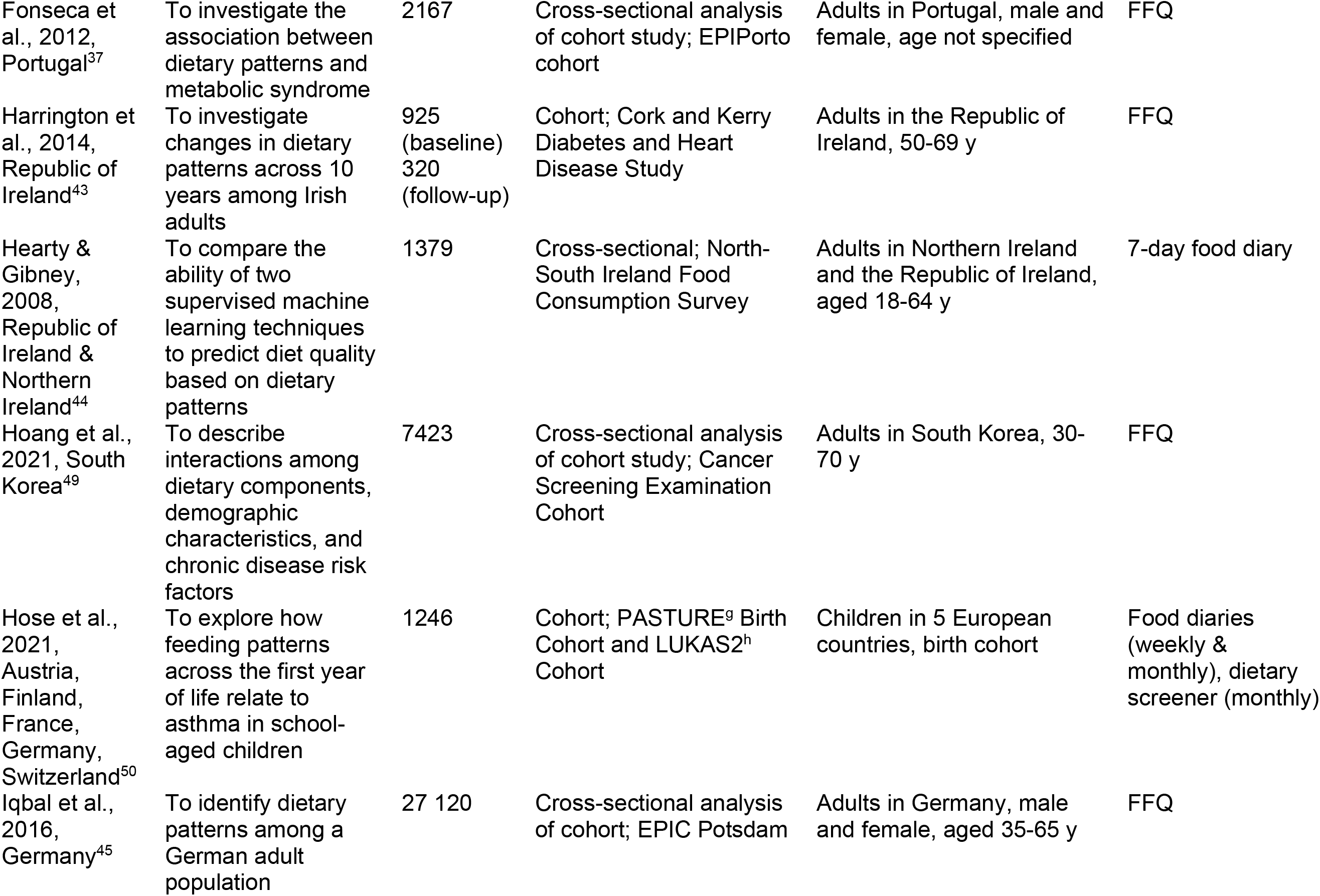
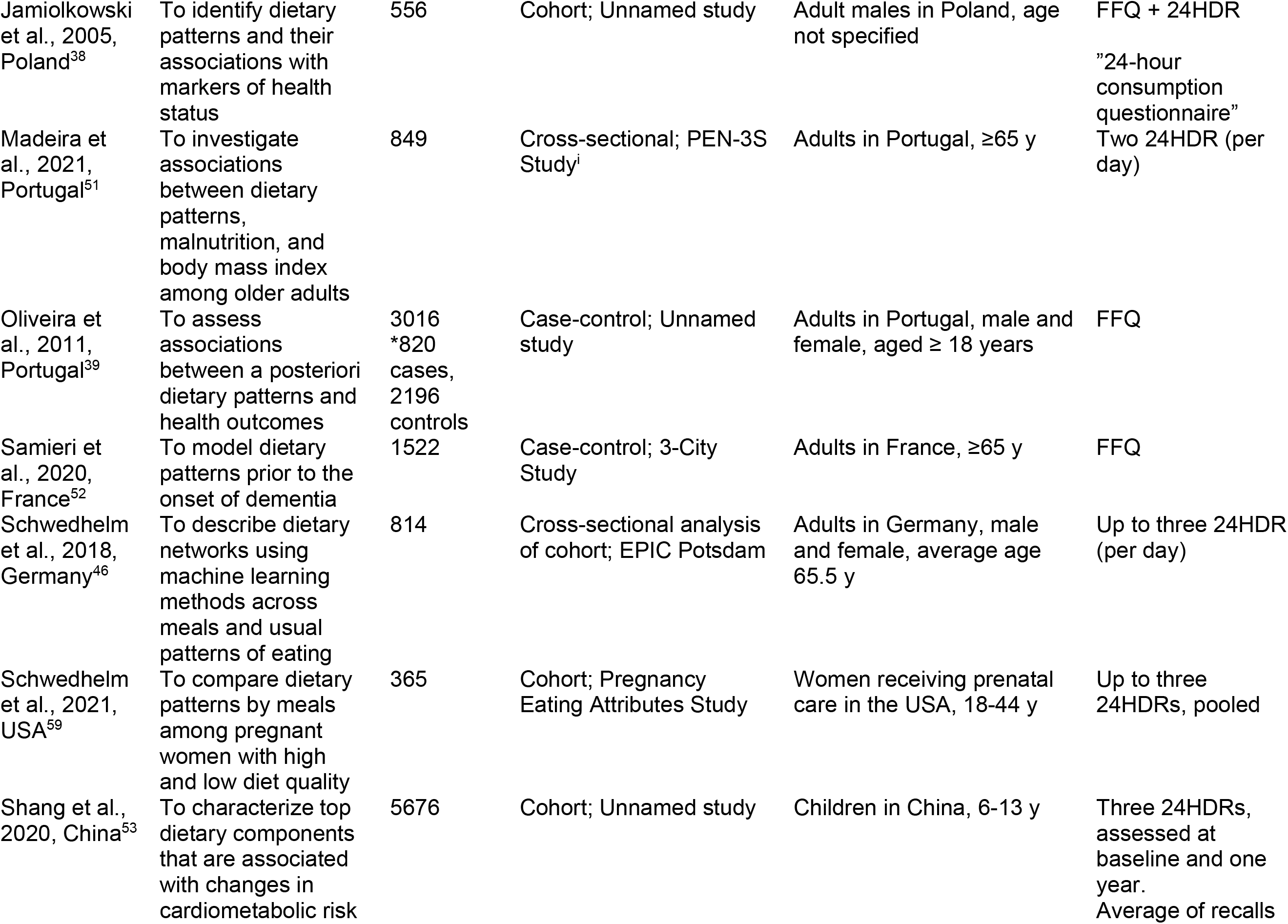
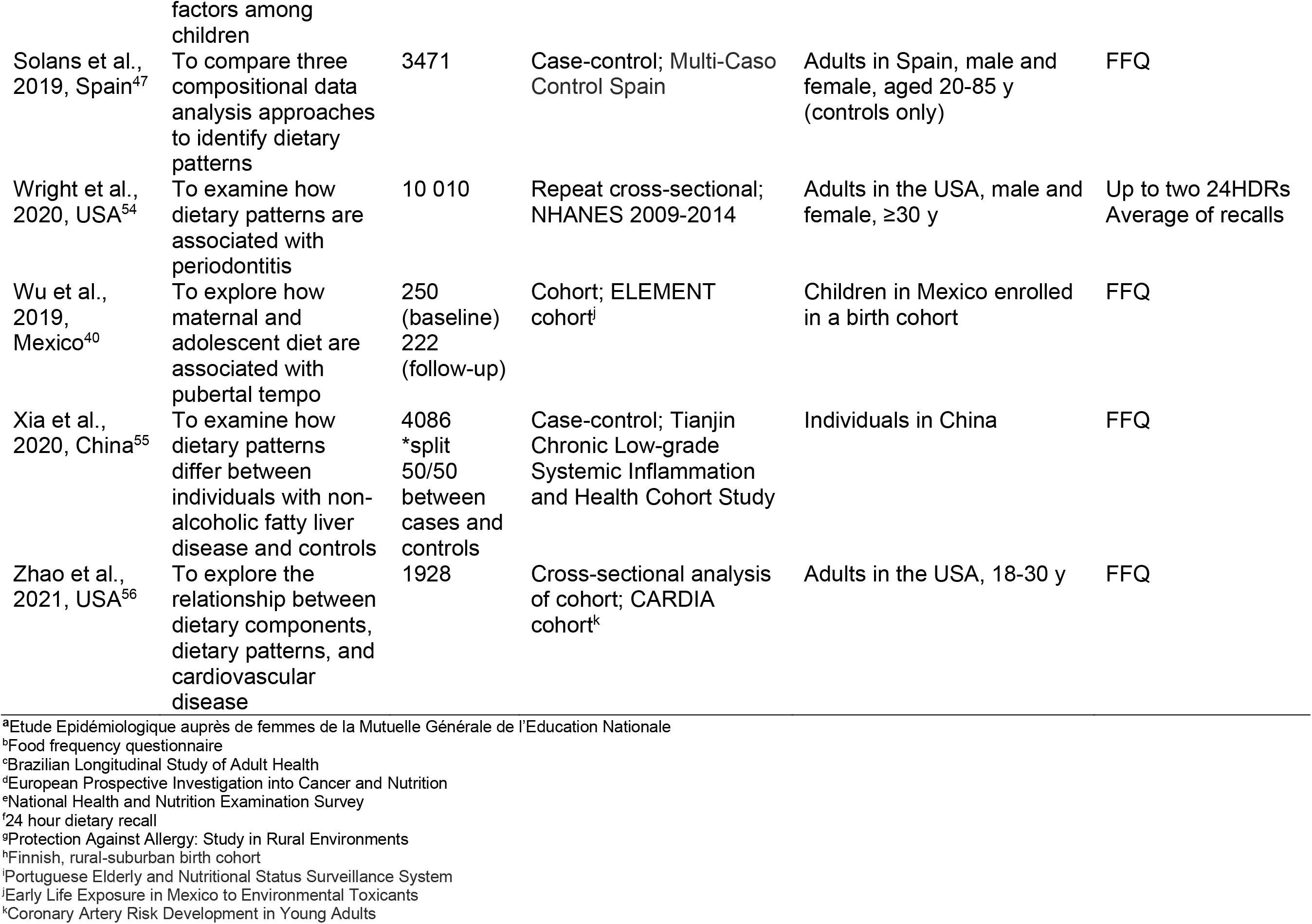
Characteristics of studies (n=24) identified in a scoping review of novel analytic methods to characterize dietary patterns.

The majority (n=15) of studies used food frequency questionnaires to assess dietary intake.^36,37,39–43,45,47–49,52,55–57^ Six studies used 24-hour recalls,^46,51,53,54,58,59^ two studies used food records/diaries,^44,50^ and one study used a food frequency questionnaire and a 24-hour recall.^38^ Among the studies using 24-hour recalls and records/diaries, one used data from a single recall that was combined with data from a food frequency questionnaire.^38^ The remaining studies, including records or recalls, averaged or combined data from two or more days of intake. Apart from averaging recalls or records, none of the included studies applied substantial efforts to mitigate measurement error present in dietary intake data. Several studies noted potential misreporting as a limitation, and only five studies specifically noted that findings may have been influenced by measurement error present in self-reported dietary assessment instruments.^40,43,51,52,59^

### Novel methods applied to identify dietary patterns

The type of methods used and how they were implemented to identify dietary patterns varied widely (**Table 3**). Nine studies applied approaches that have applications in machine learning, including classification models, neural networks, and probabilistic graphical models (**Table 4**).^36,38,44–46,49,53,56,59^ The earliest study included in this review was published in 2005 and applied neural networks to characterize dietary patterns.^38^ Fifteen studies applied other novel methods, including latent class analysis, mutual information, and treelet transform.^37,39–43,47,48,50–52,54,55,57,58^ Two studies identified dietary patterns using more than one novel method.^44,53^ Five studies included comparisons of different novel methods, though these were typically versions of the same model.^44,45,47,49,53^ For example, Solans et al. compared three models for compositional data analysis and reported that the best-performing model incorporated both investigator– and data-driven methods.^47^

**Table 3:**
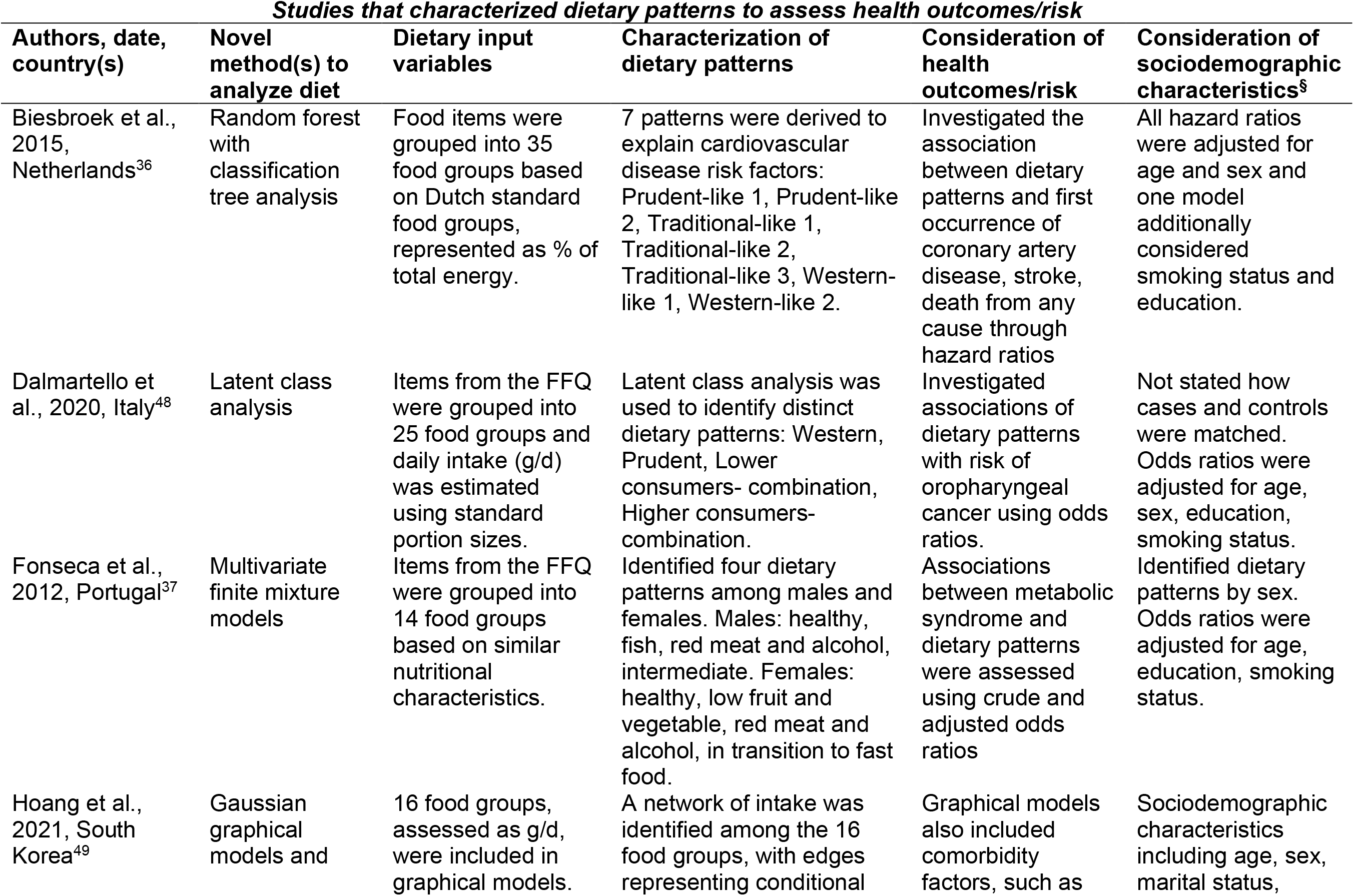
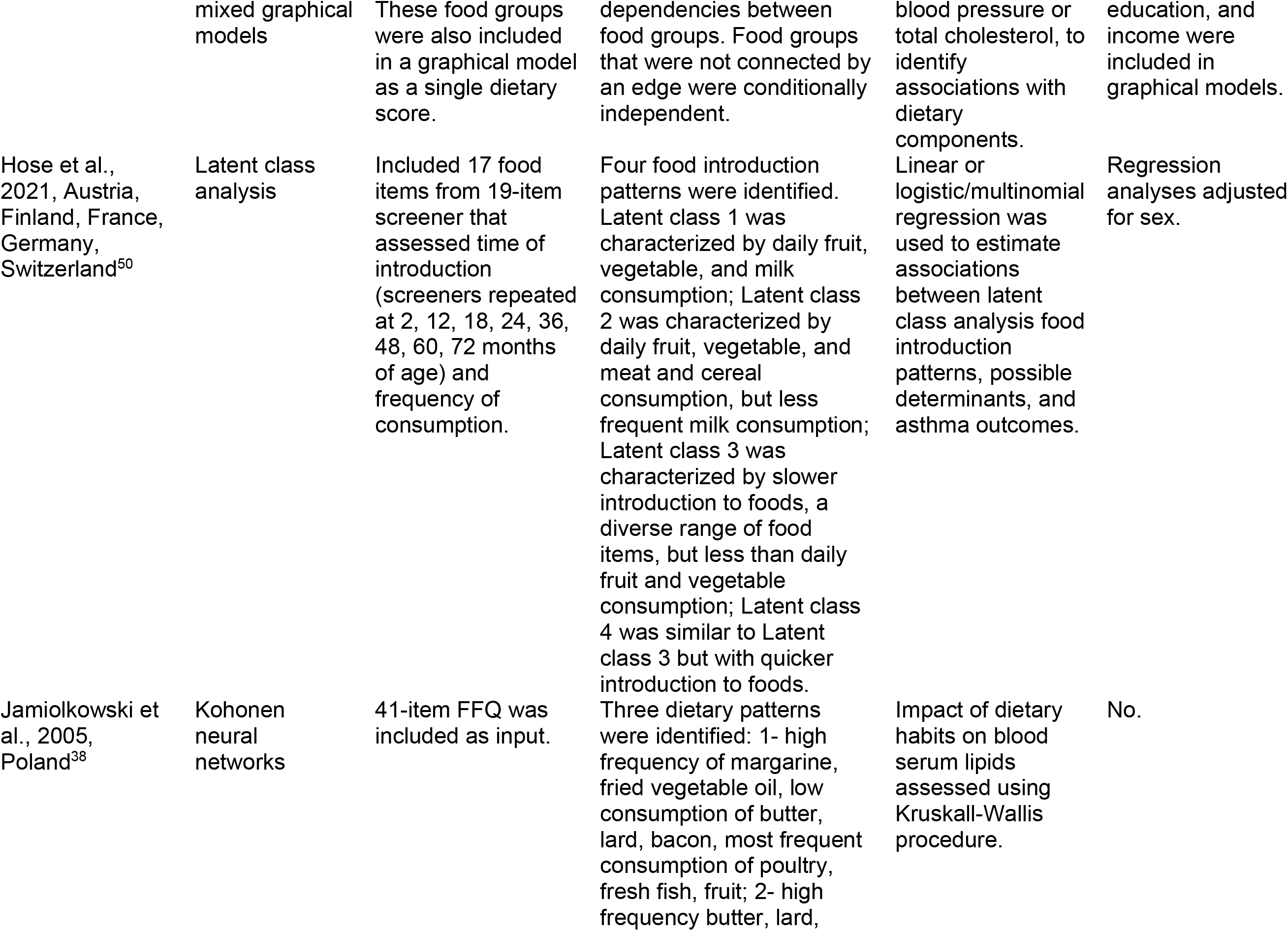
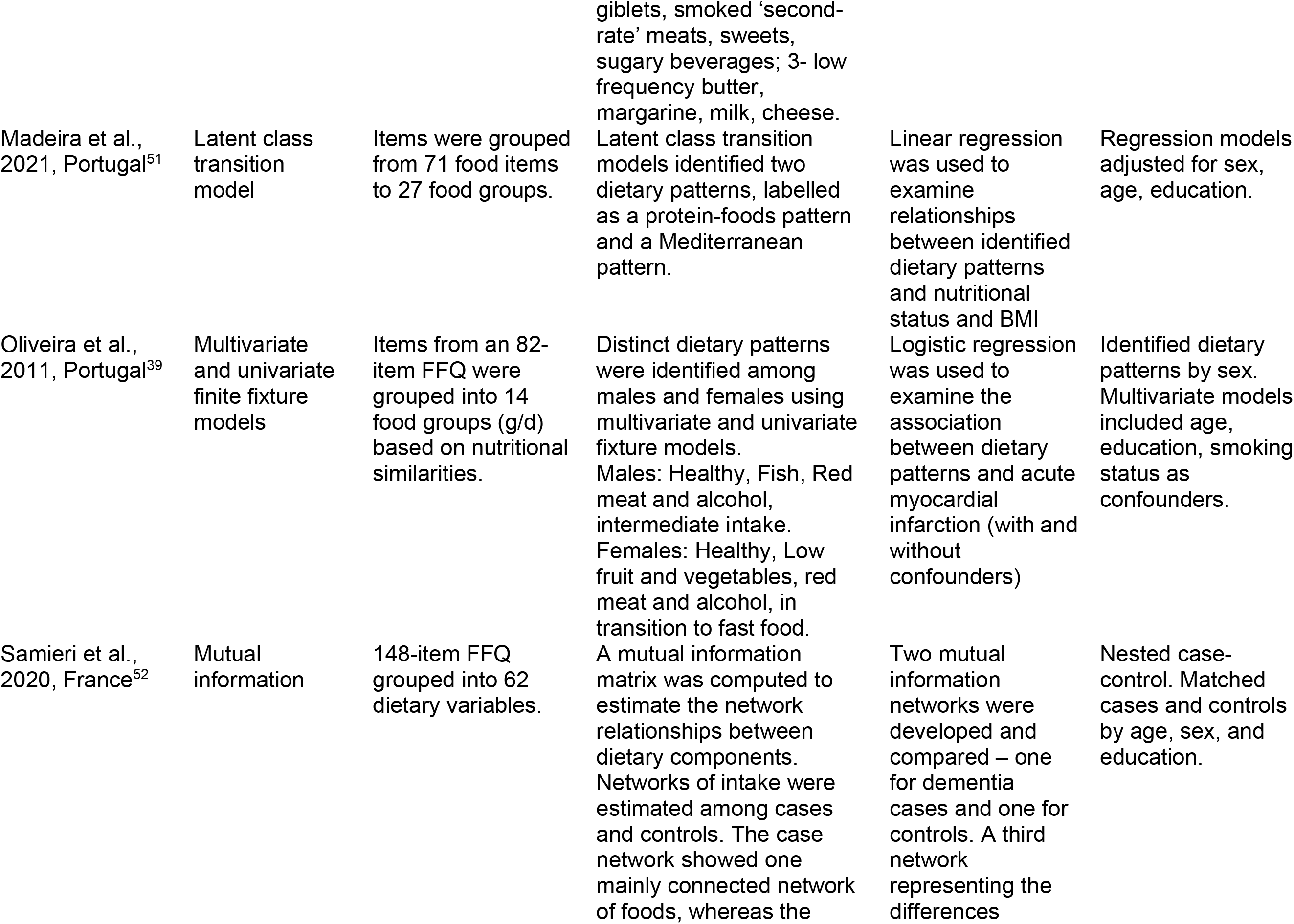
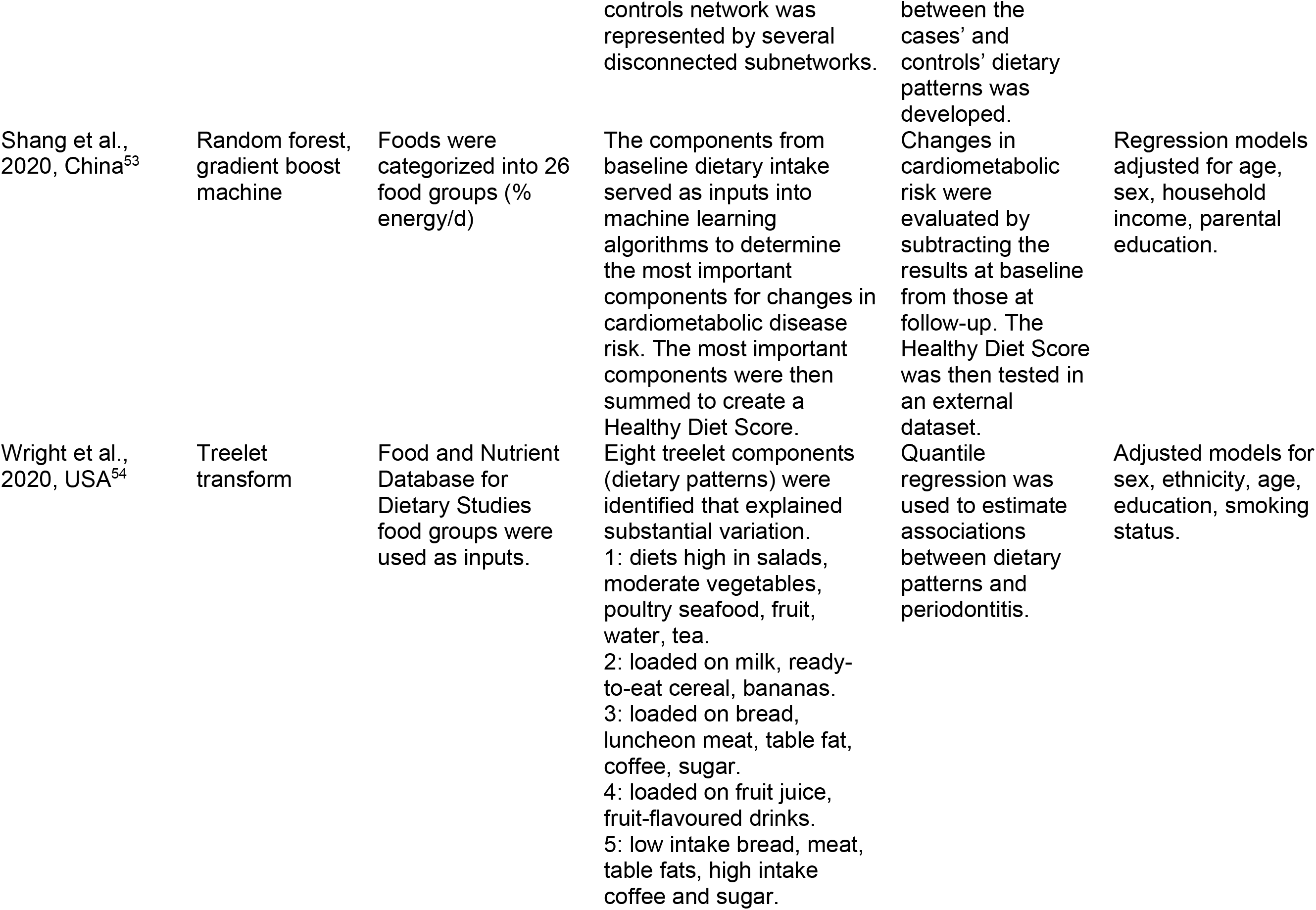
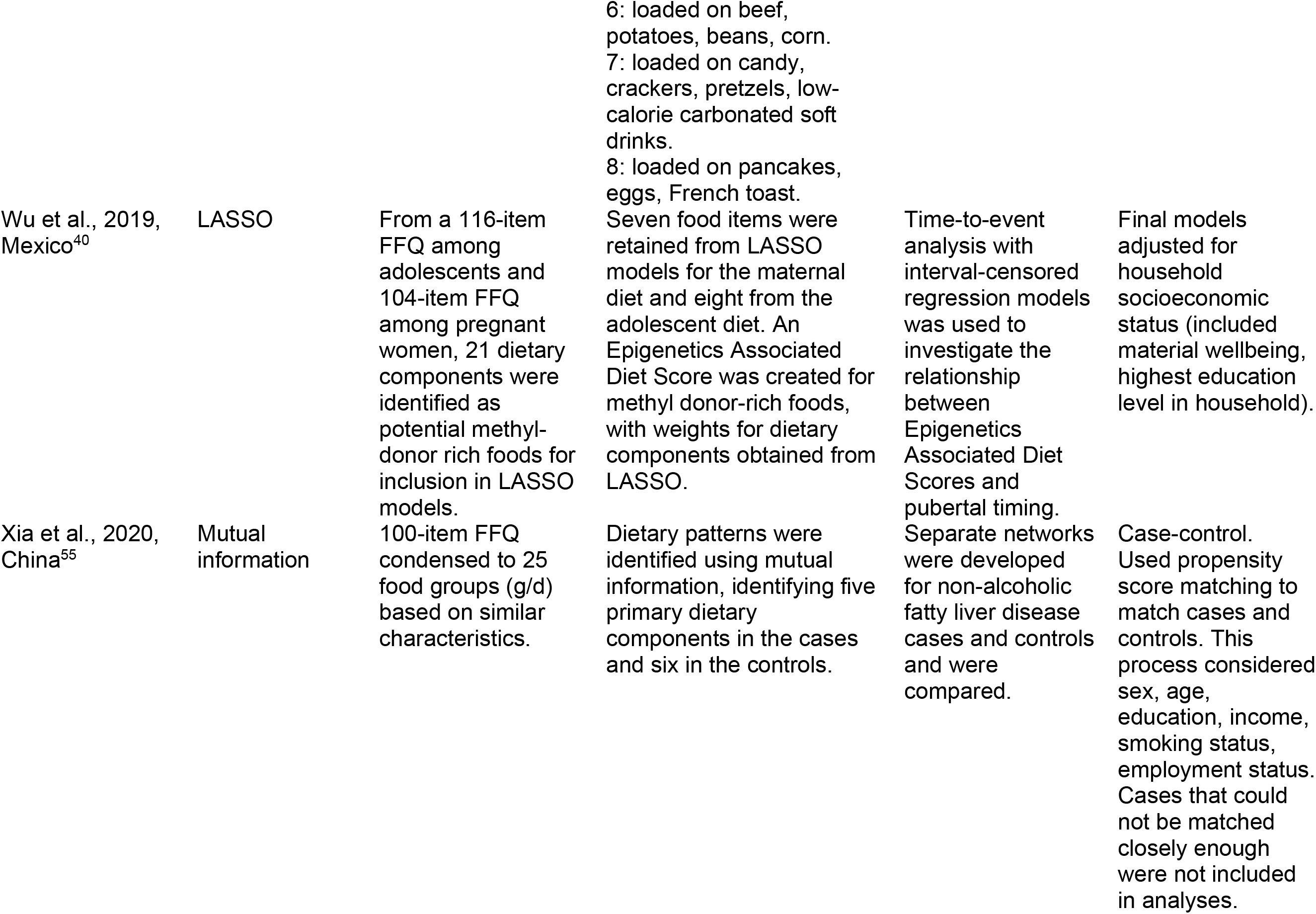
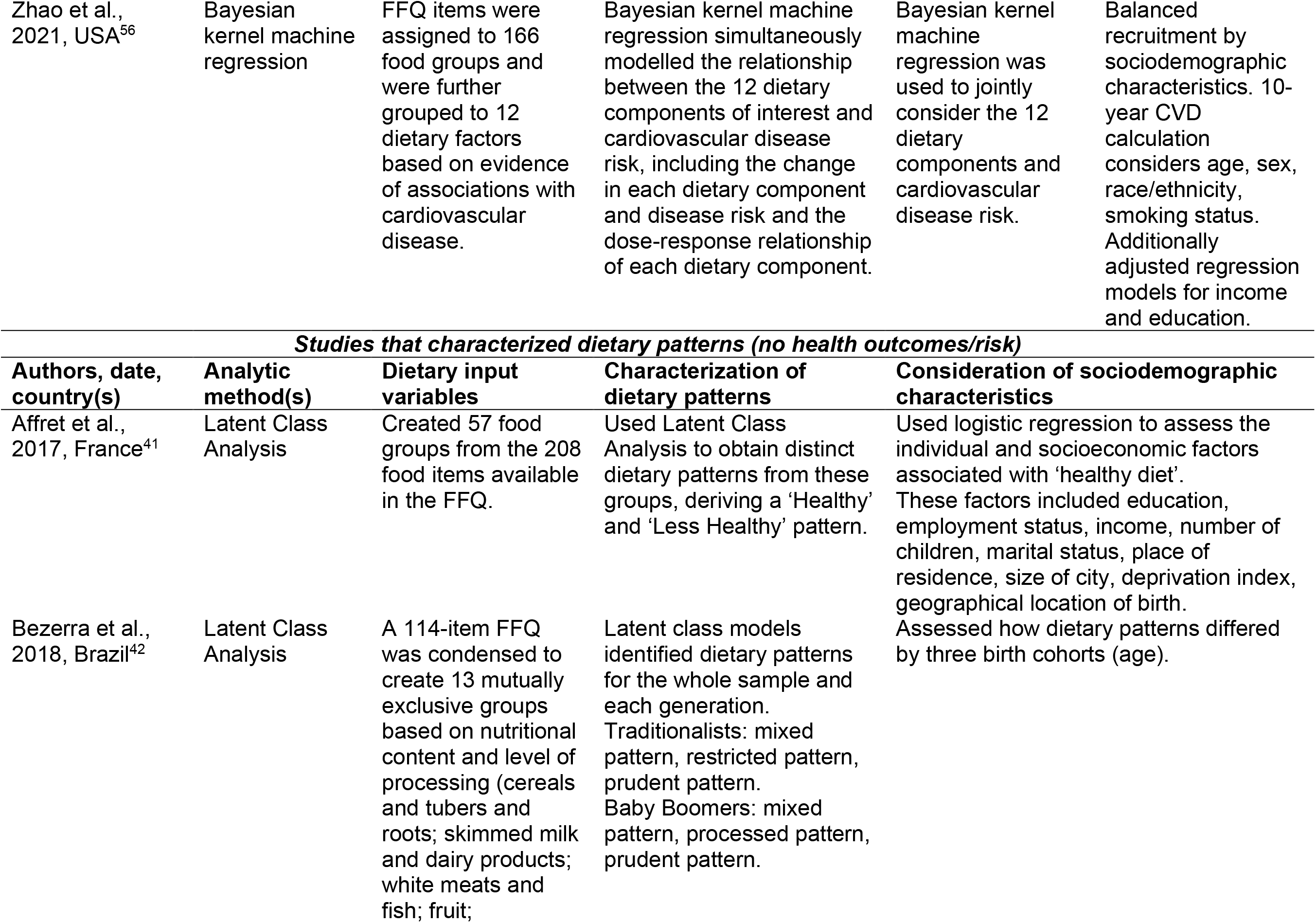
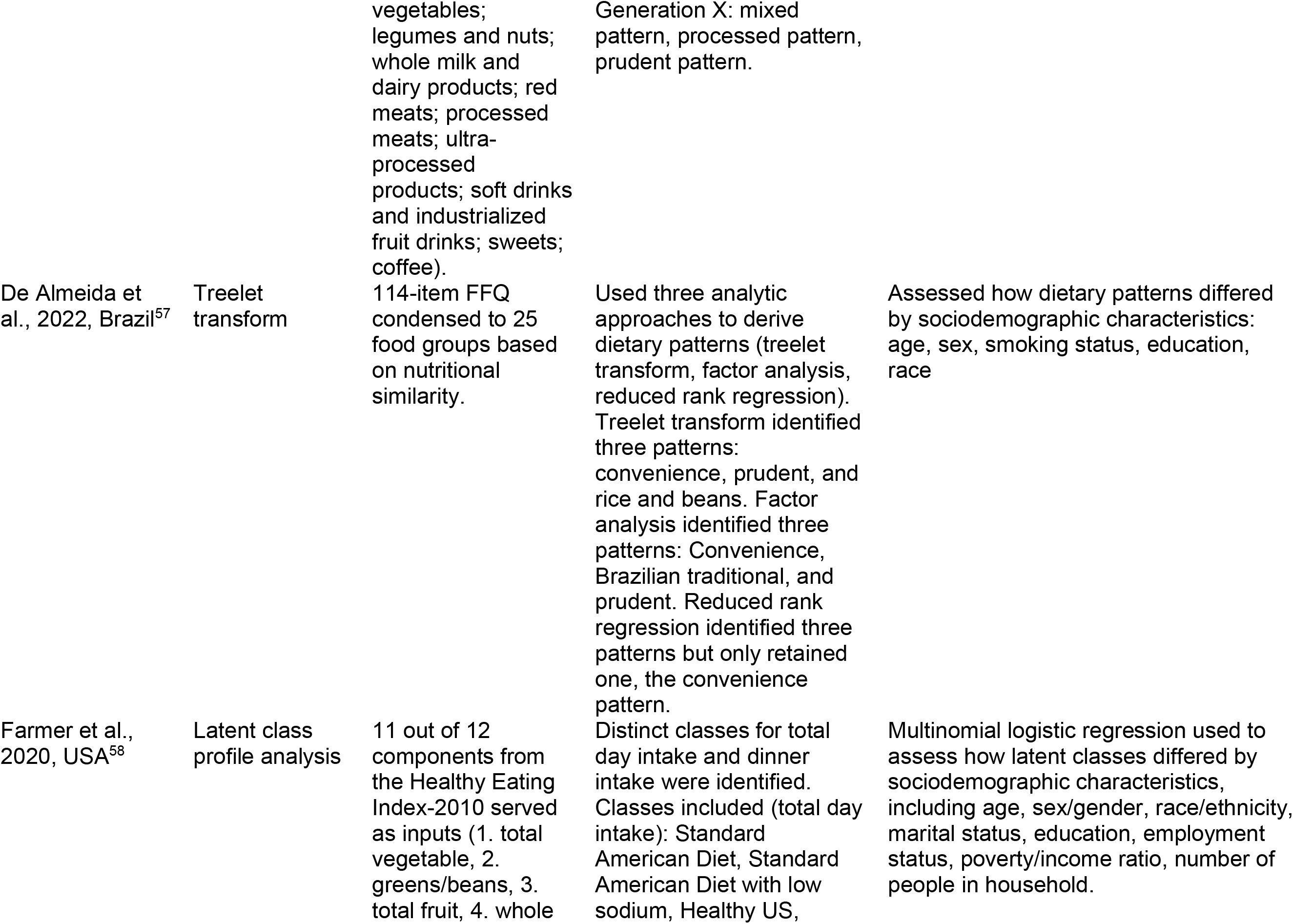
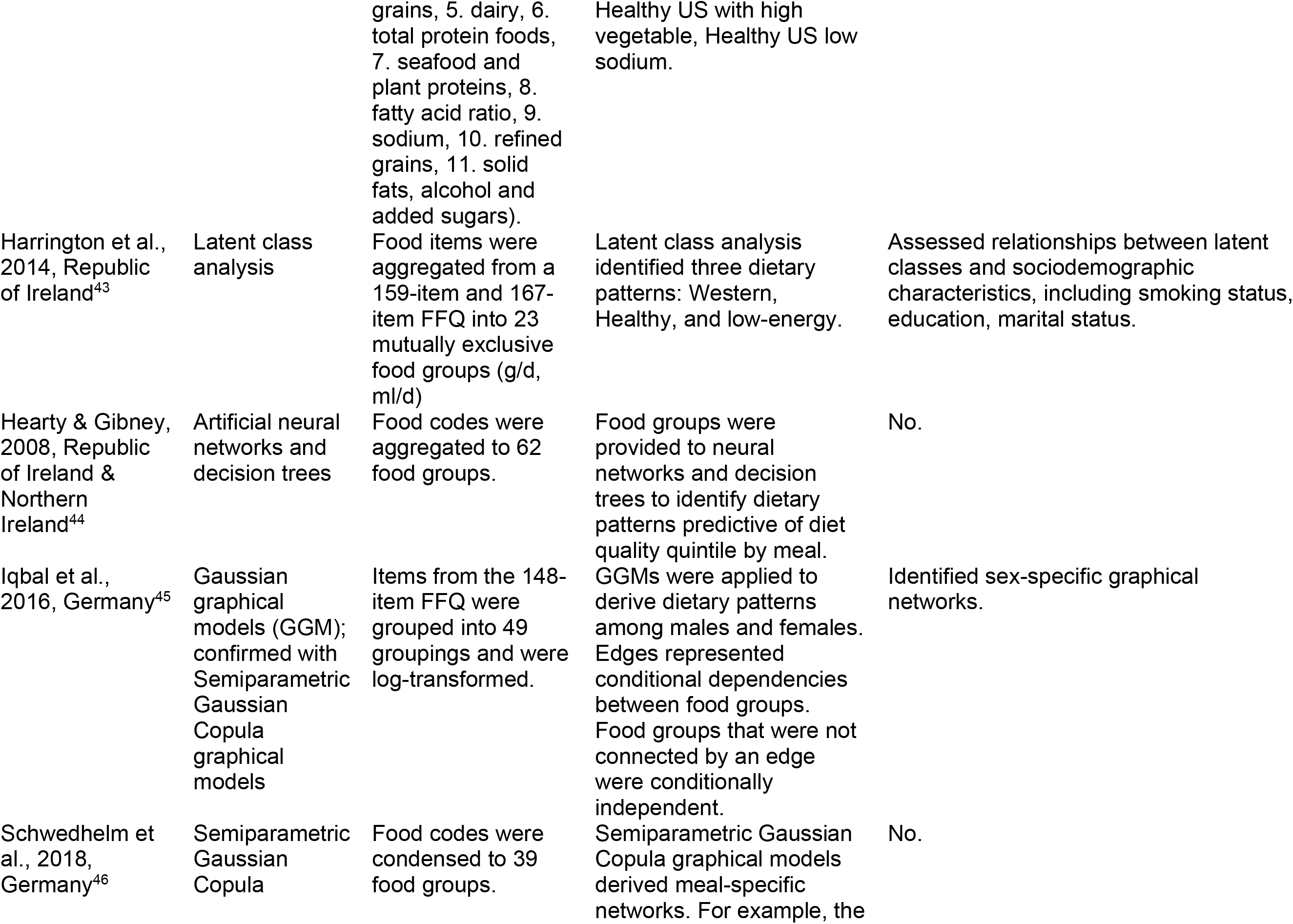
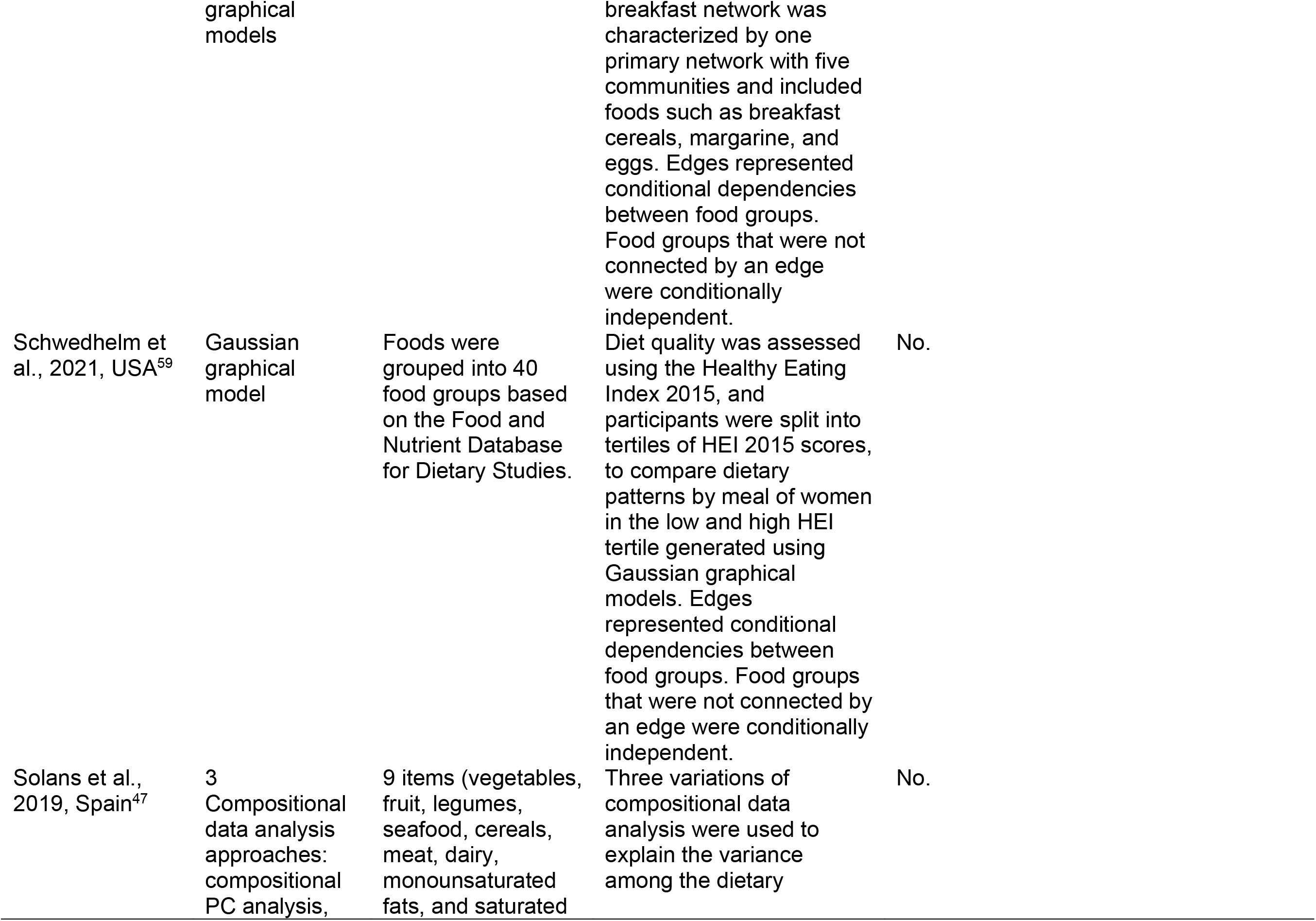
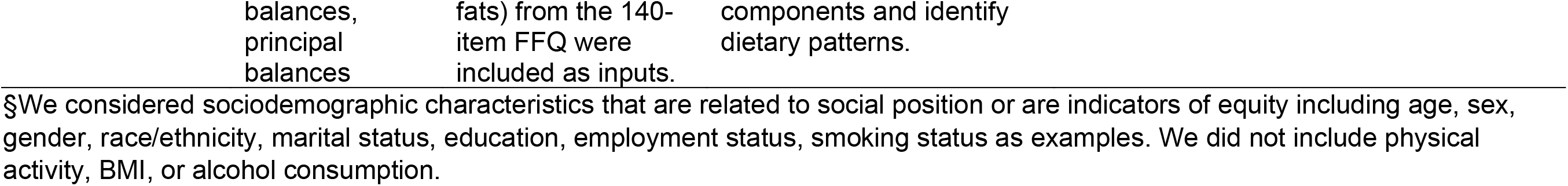
Description of dietary patterns (n=24) identified in a scoping review of novel analytic methods to characterize dietary patterns.

**Table 4:** Novel methods applied to identify dietary patterns across included studies.

| Method of analysis | Description | References |
| --- | --- | --- |
| Methods with applications in machine learning |  |  |
| Classification models | Algorithms that can be used for prediction and classification.<sup>76,77</sup> |  |
| Random forest | The random forest is generated from a wide array of trees that are randomly sampled and averaged. Random forest models can additionally be used on regressors.<sup>76</sup> | (Biesbroek et al. 2015; Shang et al. 2020) |
| Decision tree | Trees include nodes (variables) and branches, representing possible outcomes from a node. This forms an 'inverted tree', beginning with a root node and branching to many possible end nodes.<sup>77</sup> | (Hearty and Gibney 2008) |
| Neural network | The structure of this algorithm was based on the human brain and is well-suited to complex, non-linear problems.<sup>78</sup> |  |
| Artificial neural network | The algorithm starts with input nodes, which communicate, forming hidden layers of connections within the network, and provide learned output.<sup>78</sup> | (Hearty and Gibney 2008) |
| Kohonen neural network | A neural network comprised of a single layer. Data enter through input nodes and are organized to a simple structure of output nodes.<sup>79</sup> | (Jamiolkowski, Szpak, and Pawłowska 2005) |
| Probabilistic graphical models | Algorithm that can be used to determine joint relationships between nodes, forming a graphical network, where connections between nodes are represented by edges. These connections may be full- or partial-order correlations.<sup>80–82</sup> |  |
| Gaussian graphical models | Probabilistic graphical model where data follows a Gaussian distribution.<sup>81</sup> | (Hoang, Lee, and Kim 2021; Iqbal et al. 2016; Schwedhelm et al. 2021) |
| Mixed graphical models | Probabilistic graphical model where data is mixed (i.e., Gaussian and categorical).<sup>81</sup> | (Hoang, Lee, and Kim 2021) |
| Semiparametric Gaussian Copula graphical models | Probabilistic graphical model where the data is not required to be normally distributed.<sup>83</sup> | (Iqbal et al. 2016; Schwedhelm et al. 2018) |
| Other machine learning approaches |  |  |
| Gradient boost machine | Machine learning algorithm that applies a boosting ensemble method to decision trees. This allows a weak learning algorithm to provide predictions through weighted averages of weak predictions.<sup>53,84</sup> | (Shang et al. 2020) |
| Bayesian kernel machine regression | Machine learning algorithm that uses a kernel function to consider the effects of multiple exposures, covariates, and confounders on an outcome.<sup>85</sup> | (Zhao et al. 2021) |
| Compositional data analysis | Method that identifies the relative magnitude of differences between variables to determine relative importance.<sup>86,87</sup> | (Solans et al. 2019) |
| Least Absolute Shrinkage and Selection Operator (LASSO) | A method that selects the most important variables and reduces the number of variables in the model.<sup>88</sup> | (Wu et al. 2019) |
| Multivariate finite mixture models |  |  |
| Latent Class Analysis | A method that creates mutually exclusive groups (classes) of participants based on similar latent characteristics.<sup>41,42</sup> | (Affret et al. 2017; Bezerra et al. 2018; Dalmartello et al. 2020; Harrington et al. 2014; Hose et al. 2021) |
| Latent class transition model | Allows for transition between identified groups.<sup>51,89</sup> | (Madeira et al. 2021) |
| Latent class profile analysis | Latent class analysis when continuous variables are used.<sup>58</sup> | (Farmer et al. 2020) |
| Multivariate finite mixture models | Identifies mutually exclusive groupings where cluster shape is not limited.<sup>90</sup> | (Fonseca et al. 2012; Oliveira et al. 2011) |
| Mutual information | This method creates networks, estimating the amount that uncertainty is reduced in one variable by knowing another.<sup>91</sup> | (Samieri et al. 2020; Xia et al. 2020) |
| Treelet transform | Data-driven approach that reduces dimensionality and is a hybrid between hierarchical clustering and principal component analysis.<sup>92,93</sup> | (de Almeida Alves et al. 2022; Wright et al. 2020) |

\*Studies may have used more than one novel approach to characterize dietary patterns.

In twelve studies, two to eight distinct dietary patterns, such as the ‘prudent’ pattern or ‘Western’ pattern, were identified using methods such as latent class analysis, treelet transform, random forest with classification tree analysis, and multivariate finite mixture models.^36–38,41–43,48,50,51,54,57,58^ Six studies applied network methods, including probabilistic graphical models and mutual information, to identify networks of dietary patterns among populations.^45,46,49,52,55,59^

Dynamism, or how dietary patterns vary across time, was incorporated into four studies’ characterization or analysis of dietary patterns. Three studies incorporated stratification by meals to consider dynamism.^44,46,59^ In two studies using graphical models, separate networks were created for each meal to provide insights into how patterns of intake vary throughout the day.^46,59^ Hearty and Gibney used decision trees and neural networks and ran models by meals based on 62 food groups to predict diet quality.^44^ Additionally, one study considered dynamism by using ANOVA and chi-square tests to descriptively show how a variety of characteristics were associated with stable or changing dietary patterns characterized using latent class analysis.^43^

Fourteen studies examined relationships between dietary patterns characterized using novel methods and variables indicative of health risk or outcomes, such as periodontitis, cardiovascular disease, and metabolic syndrome (Table 3).^36–40,48–56^ Six studies included longitudinal analysis of the relationship between dietary patterns and health outcomes.^36,38,40,50,52,53^ Most studies that examined health risk or outcomes first identified dietary patterns using a novel method and then investigated relationships with health outcomes using regression models.^36–40,48,50,51,54^ In contrast, some studies incorporated variables indicative of health outcomes or risk directly into the machine learning models.^49,56^ For example, Zhao et al.^56^ applied Bayesian kernel machine regression, a machine learning model designed to incorporate high dimensional data, to jointly model the relationship between several dietary components and cardiovascular disease risk. Similarly, a paper by Hoang et al.^49^ included health variables within mixed graphical models, though directionality of diet-health relationships could not be ascertained given the cross-sectional nature of the data. In two case-control studies, dietary patterns were identified using mutual information to estimate dietary pattern networks, with stratification by health outcomes.^52,55^

Nineteen studies considered sociodemographic characteristics, such as sex, age, race/ethnicity, education, and income.^36,37,39–43,45,48–58^ In one case, sociodemographic characteristics were included in models used to characterize dietary patterns.^49^ Two studies stratified by sociodemographic characteristics, examining dietary patterns by sex^45^ or age groups.^42^ Studies that used case-control designs typically considered sociodemographic characteristics through matching.^52,55^ In the remaining studies that considered sociodemographic characteristics, these were incorporated in regression models to explore how dietary patterns characterized using novel methods were associated with health and other characteristics.

Two studies included comparisons of novel methods and traditional statistical approaches.^36,57^ While there was overlap between the patterns identified through novel and traditional approaches in these studies, the patterns identified differed across approaches. For instance, Biesbroek et al.^36^ found that dietary patterns identified through reduced rank regression were more strongly associated with coronary artery disease compared to those identified through random forest with classification tree analysis.

## Discussion

The application of novel methods to dietary pattern research is rapidly expanding, with the aim of better understanding their complexity and how they are related to health and other factors. Many studies used methods that characterize distinct dietary patterns based on the population being studied, such as the ‘prudent’ pattern or the ‘Western’ pattern. Most studies used cross-sectional data, limiting opportunities to examine the effect of dietary patterns on health.

Methods newly being applied in this field offer promising capacity to better understand the totality of dietary patterns and synergistic relationships among dietary components when compared with traditional approaches that do not assume synergy.^4,14^ Given the large variation in how dietary patterns were characterized using novel methods, multidimensionality and potential synergistic relationships between dietary components were considered and presented in a range of ways, from latent classes to networks. Several studies incorporated dynamism into their consideration of dietary patterns, though in most cases this was through stratification, for example, by meal, rather than through direct use of novel methods.^43,44,46,59^ In these cases, it was a combination of input variables, stratification by time, and the novel method that enabled explorations of dynamism.

The methods highlighted have a range of strengths and limitations for the characterization of dietary patterns. Methods that focused on the classification of distinct patterns allowed for the assessment of relationships between these patterns and health outcomes or other indicators of interest but explored the interrelationships between dietary components to a lesser degree.^36–38,41–43,48,50,51,54,57,58^ Other methods were designed to better consider synergistic relationships among dietary components, such as compositional data analysis, mutual information, or probabilistic graphical models, but required further analyses, such as the development of a score, to assess relationships with health outcomes.^45–47,49,52,55,59^

There are trade-offs between novel and traditional methods that should be considered when contemplating the most appropriate methods for a given study. Though potential benefits such as a greater ability to discern multidimensionality may be desirable, these must be weighed against the implications for interpretability and computational costs. The application of novel methods may not always yield insights beyond those gained from traditional approaches. For example, Biesbroek et al.^36^ found that random forest models did not outperform reduced rank regression when examining associations of dietary patterns with coronary artery disease. Conversely, a study that was not included in this review because it first identified dietary patterns using a traditional method—principal component analysis—found that machine learning algorithms were better able to classify the identified dietary patterns according to cardiometabolic risk compared to traditional approaches.^60^

Several sociodemographic characteristics are indicators of systemic health inequity and have been shown to be associated with dietary patterns among populations.^61–63^ The degree to which studies incorporated sociodemographic characteristics into their consideration of dietary patterns or relationships between dietary patterns and health varied, with adjusted regression models applied after dietary patterns were characterized as the most common approach. Consistent with nutrition research more broadly,^63–65^ there was little consideration of possible interactions among sociodemographic characteristics in relation to dietary patterns. Methods particularly suited to pattern recognition and complexity could be leveraged to simultaneously explore potential joint relationships among facets of social identity and dietary patterns^66^ and advance our understanding of how broader systems of oppression and intersecting characteristics contribute to dietary patterns.^62^

Beyond the inclusion of sociodemographic characteristics in models, considering equity from the beginning of study design is a critical consideration given potential bias in data and algorithms that can have immense implications for those who already experience inequities, such as structural racism.^67–69^ The included studies did not explicitly discuss the incorporation of equity into study design, and many conducted secondary analyses of existing datasets. The use of directed acyclic graphs has been identified as a potential solution to mitigate some possible issues with bias through careful model design^67^ and has been applied in other domains of nutrition research using novel methods.^14^ Engaging individuals with lived experience and the integration of interdisciplinary teams with broad expertise that can combine content knowledge with data-driven approaches can help to mitigate potential bias in algorithms.^66^

The level of description of methods varied and it was sometimes challenging to decipher the specifics of how novel methods were applied. Although the Strengthening the Reporting of Observational Studies in Epidemiology—Nutritional Epidemiology (STROBE-nut) reporting guidelines provide guidance for transparently reporting nutritional epidemiology and dietary assessment research,^70^ it was not designed specifically for the methods used in the studies considered in this review and the ways in which they are being applied in dietary patterns research. Other reporting guidelines, such as the Consolidated Standards of Reporting Trials (CONSORT), have been extended to consider the application of artificial intelligence (AI).^71^ Motivations related to the extension of CONSORT included inadequate reporting of studies using AI and the lack of full consideration of potential sources of bias specific to AI within existing reporting guidelines.^71^ Relevant items added to CONSORT-AI pertain to the role of AI in the study, the nature of the data used in AI systems, and how humans interacted with AI systems, for example.^71^ The extension of reporting guidelines such as STROBE-nut to consider applications of AI, including machine learning, and other methods that are becoming more commonly used, may facilitate consistent and complete reporting and improved comparability of studies. Reporting guidelines should continue to emphasize strategies applied to mitigate measurement error in dietary intake data,^70^ as studies using novel methods are not immune to the effects of error on findings.^72^ Along with reporting guidelines, the development of tailored quality appraisal tools may facilitate synthesis of high-quality evidence to inform recommendations about dietary patterns and health.

This review provides a snapshot of a rapidly evolving field,^73,74^ with the involvement of an interdisciplinary team of researchers lending to a robust consideration of emerging methods in dietary patterns research. While prior reviews have provided perspectives on the potential applications of machine learning within the field of nutrition,^19–21^ this review considered dietary patterns in particular, as well as considering approaches beyond machine learning that have not been traditionally used in this area, broadening the scope compared to prior reviews.^22,75^ The search terms were informed by preliminary searching, though it is unlikely that all relevant articles applying novel methods to characterize dietary patterns were captured. This is partially driven by the wide range of descriptors used for these methods and the lack of reporting standards. As well, determining whether a method is novel is somewhat subjective. Methods such as factor analysis and principal component analysis once revolutionized dietary pattern analysis, providing data-driven approaches to identify patterns.^15^ Now, they are widely applied and recognized as limited in their capabilities to capture complexity compared to some newer approaches. Further, the search terms skewed toward multidimensionality versus dynamism, potentially overlooking some studies focusing on variation of dietary patterns over time or across eating occasions. Nonetheless, this review documents an acceleration of the application of a range of novel methods to dietary patterns research and captures a broad scope of methods being used to characterize these patterns, highlighting the need for researchers to develop the lexicon and knowledge needed to interpret the emerging literature.

## Conclusion

The findings of this review indicate a strong motivation to apply novel methods, including but not limited to machine learning, to improve understanding of dietary patterns and how they relate to health and other factors. The application of these methods may help us to learn about complex relationships that may not be possible to discern through traditional approaches. However, these methods may not be suitable for every question and do not necessarily overcome the limitations of more traditional approaches.

Given the proliferation of these methods, it is becoming increasingly worthwhile for nutrition researchers to have at least a basic understanding of novel methods such as machine learning and latent class analysis, so they can interpret the results of emerging studies. The development and implementation of reporting guidelines and quality appraisal mechanisms for studies that apply novel methods may improve the capacity for synthesis of evidence generated to inform strategies that promote improved population health and well-being.

## Data Availability

Extracted metadata for all articles is available upon reasonable request to the authors.

## Acknowledgements

We thank research librarian Jackie Stapleton (JS) of the University of Waterloo for support with the search strategy.

## Contributions

SIK conceived of the review and planned it with the co-authors; AR, AP, SH, SIK conducted the search and screening; JMH and TEW conducted extraction; LA conducted verification; JMH led the first draft of the manuscript with support from AP to write the methods; all co-authors provided critical input to the manuscript and all co-authors read and approved the final manuscript.

## References

1. Afshin A, Sur PJ, Fay KA, et al. Health effects of dietary risks in 195 countries, 1990–2017: a systematic analysis for the Global Burden of Disease Study 2017. The Lancet. 2019;393(10184):1958–1972. doi:10.1016/S0140-6736(19)30041-8

2. English LK, Ard JD, Bailey RL, et al. Evaluation of dietary patterns and all-cause mortality: A systematic review. JAMA network open. 2021;4(8). doi:10.1001/JAMANETWORKOPEN.2021.22277

3. Mozaffarian D, Rosenberg I, Uauy R. History of modern nutrition science—implications for current research, dietary guidelines, and food policy. BMJ. 2018;361. doi:10.1136/BMJ.K2392

4. Reedy J, Subar AF, George SM, Krebs-Smith SM. Extending methods in dietary patterns research. Nutrients. 2018;10(5). doi:10.3390/NU10050571

5. Schulz CA, Oluwagbemigun K, Nöthlings U. Advances in dietary pattern analysis in nutritional epidemiology. Eur J Nutr. 2021;60(8):4115–4130. doi:10.1007/s00394-021-02545-9

6. Herforth A, Arimond M, Álvarez-Sánchez C, Coates J, Christianson K, Muehlhoff E. A global review of food-based dietary guidelines. Advances in Nutrition. 2019;10(4):590. doi:10.1093/ADVANCES/NMY130

7. National Academies of Sciences, Engineering, and Medicine; Health and Medicine Division; Food and Nutrition Board; Committee to Review the Process to Update the Dietary Guidelines for Americans. Redesigning the Process for Establishing the Dietary Guidelines for Americans. National Academies Press; 2017. doi:10.17226/24883

8. Imamura F, Micha R, Khatibzadeh S, et al. Dietary quality among men and women in 187 countries in 1990 and 2010: a systematic assessment. The Lancet Global Health. 2015;3(3):e132–e142. doi:10.1016/S2214-109X(14)70381-X

9. Delormier T, Frohlich K, Potvin L. Food and eating as social practice – understanding eating patterns as social phenomena and implications for public health. Sociolgy of Health & Illness. 2009;31(2):215–228. doi:10.1111/j.1467-9566.2008.01128.x

10. Ocké MC. Evaluation of methodologies for assessing the overall diet: dietary quality scores and dietary pattern analysis. Proceedings of the Nutrition Society. 2013;72(2):191–199. doi:10.1017/S0029665113000013

11. Shams-White MM, Pannucci TE, Lerman JL, et al. Healthy Eating Index-2020: Review and update process to reflect the Dietary Guidelines for Americans, 2020-2025. J Acad Nutr Diet. 2023;123(9):1280–1288. doi:10.1016/j.jand.2023.05.015

12. Brassard D, Munene LAE, Pierre SS, et al. Development of the Healthy Eating Food Index (HEFI)-2019 measuring adherence to Canada’s Food Guide 2019 recommendations on healthy food choices. *Applied Physiology*, Nutrition, and Metabolism. 2022;47(5):595–610. doi:10.1139/APNM-2021-0415

13. Kirkpatrick SI, Reedy J, Krebs-Smith SM, et al. Applications of the Healthy Eating Index for surveillance, epidemiology, and intervention research: Considerations and caveats. Journal of the Academy of Nutrition and Dietetics. 2018;118(9):1603. doi:10.1016/J.JAND.2018.05.020

14. Bodnar LM, Cartus AR, Kirkpatrick SI, et al. Machine learning as a strategy to account for dietary synergy: an illustration based on dietary intake and adverse pregnancy outcomes. The American Journal of Clinical Nutrition. 2020;111(6):1235–1243. doi:10.1093/AJCN/NQAA027

15. Hu FB. Dietary pattern analysis: a new direction in nutritional epidemiology. Current Opinion in Lipidology. 2002;13(1):3–9. doi:10.1097/00041433-200202000-00002

16. Michels KB, Schulze MB. Can dietary patterns help us detect diet-disease associations? Nutr Res Rev. 2005;18(2):241–248. doi:10.1079/NRR2005107

17. Reedy J, Krebs-Smith SM, Hammond RA, Hennessy E. Advancing the science of dietary patterns research to leverage a complex systems approach. Journal of the Academy of Nutrition and Dietetics. 2017;117(7):1019–1022. doi:10.1016/J.JAND.2017.03.008

18. National Academies of Sciences, Engineering, and Medicine; Health and Medicine Division; Food and Nutrition Board; Alice Vorosmarti and Joe Alper, Rapporteurs. The Role of Advanced Computation, Predictive Technologies, and Big Data Analytics in Food and Nutrition Research. National Academies Press; 2024. Accessed May 10, 2024. https://www.nationalacademies.org/our-work/the-role-of-advanced-computation-predictive-technologies-and-big-data-analytics-in-research-related-to-food-and-nutrition-a-workshop-series

19. Kirk D, Kok E, Tufano M, Tekinerdogan B, Feskens EJM, Camps G. Machine learning in nutrition research. Advances in Nutrition. 2022;13(6):2573–2589. doi:10.1093/ADVANCES/NMAC103

20. Morgenstern JD, Rosella LC, Costa AP, Souza RJ de, Anderson LN. Perspective: Big data and machine learning could help advance nutritional epidemiology. Advances in Nutrition. 2021;12(3):621–631. doi:10.1093/ADVANCES/NMAA183

21. Côté M, Lamarche B. Artificial intelligence in nutrition research: perspectives on current and future applications. Appl Physiol Nutr Metab. Published online 2021:1–8. doi:10.1139/apnm-2021-0448

22. Oliveira Chaves L, Gomes Domingos AL, Louzada Fernandes D, Ribeiro Cerqueira F, Siqueira-Batista R, Bressan J. Applicability of machine learning techniques in food intake assessment: A systematic review. Critical Reviews in Food Science and Nutrition. 2023;63(7):902–919. doi:10.1080/10408398.2021.1956425

23. Le Glaz A, Haralambous Y, Kim-Dufor DH, et al. Machine learning and natural language processing in mental health: Systematic review. J Med Internet Res. 2021;23(5):e15708. doi:10.2196/15708

24. Payedimarri AB, Concina D, Portinale L, et al. Prediction Models for Public Health Containment Measures on COVID-19 Using Artificial Intelligence and Machine Learning: A Systematic Review. Int J Environ Res Public Health. 2021;18(9):4499. doi:10.3390/ijerph18094499

25. Sidey-Gibbons JAM, Sidey-Gibbons CJ. Machine learning in medicine: a practical introduction. BMC Medical Research Methodology. 2019;19(1):64. doi:10.1186/s12874-019-0681-4

26. Morgenstern JD, Buajitti E, O’Neill M, et al. Predicting population health with machine learning: a scoping review. BMJ Open. 2020;10(10):e037860. doi:10.1136/BMJOPEN-2020-037860

27. Chapter 11: Scoping reviews. In: JBI Manual for Evidence Synthesis. JBI; 2020. doi:10.46658/JBIMES-20-12

28. Arksey H, O’Malley L. Scoping studies: towards a methodological framework. International Journal of Social Research Methodology. 2005;8(1):19–32. doi:10.1080/1364557032000119616

29. Tricco AC, Lillie E, Zarin W, et al. PRISMA Extension for Scoping Reviews (PRISMA-ScR): Checklist and Explanation. Ann Intern Med. 2018;169(7):467–473. doi:10.7326/M18-0850

30. Newby PK, Tucker KL. Empirically derived eating patterns using factor or cluster analysis: A review. Nutrition Reviews. 2004;62(5):177–203. doi:10.1111/j.1753-4887.2004.tb00040.x

31. Krebs-Smith SM, Subar AF, Reedy J. Examining dietary patterns in relation to chronic disease: Matching measures and methods to questions of interest. Circulation. 2015;132(9):790–793. doi:10.1161/CIRCULATIONAHA.115.018010

32. Covidence-Better systematic review management. Covidence. Accessed September 12, 2023. https://www.covidence.org/

33. Gwet KL. Computing inter-rater reliability and its variance in the presence of high agreement. British Journal of Mathematical and Statistical Psychology. 2008;61(1):29–48. doi:10.1348/000711006X126600

34. Cicchetti DV, Feinstein AR. High agreement but low kappa: II. Resolving the paradoxes. Journal of Clinical Epidemiology. 1990;43(6):551–558. doi:10.1016/0895-4356(90)90159-M

35. Belur J, Tompson L, Thornton A, Simon M. Interrater reliability in systematic review methodology: Exploring variation in coder decision-making. Sociological Methods & Research. 2021;50(2):837–865. doi:10.1177/0049124118799372

36. Biesbroek S, van der A DL, Brosens MCC, et al. Identifying cardiovascular risk factor-related dietary patterns with reduced rank regression and random forest in the EPIC-NL cohort. Am J Clin Nutr. 2015;102(1):146–154. doi:10.3945/ajcn.114.092288

37. Fonseca MJ, Gaio R, Lopes C, Santos AC. Association between dietary patterns and metabolic syndrome in a sample of Portuguese adults. Nutr J. 2012;11:64. doi:10.1186/1475-2891-11-64

38. Jamiołkowski J, Szpak A, Pawłowska D. Dietary habits of men from Podlasie region of Poland in the years 1987-1998 analysed with self-organizing neural networks. Rocz Akad Med Bialymst. 2005;50 Suppl 1:220–224.

39. Oliveira A, Rodríguez-Artalejo F, Gaio R, Santos AC, Ramos E, Lopes C. Major habitual dietary patterns are associated with acute myocardial infarction and cardiovascular risk markers in a southern European population. J Am Diet Assoc. 2011;111(2):241–250. doi:10.1016/j.jada.2010.10.042

40. Wu Y, Sánchez BN, Goodrich JM, et al. Dietary exposures, epigenetics and pubertal tempo. Environ Epigenet. 2019;5(1):dvz002. doi:10.1093/eep/dvz002

41. Affret A, Severi G, Dow C, et al. Socio-economic factors associated with a healthy diet: results from the E3N study. Public Health Nutr. 2017;20(9):1574–1583. doi:10.1017/S1368980017000222

42. Bezerra IN, Bahamonde NMSG, Marchioni DML, et al. Generational differences in dietary pattern among Brazilian adults born between 1934 and 1975: a latent class analysis. Public Health Nutr. 2018;21(16):2929–2940. doi:10.1017/S136898001800191X

43. Harrington JM, Dahly DL, Fitzgerald AP, Gilthorpe MS, Perry IJ. Capturing changes in dietary patterns among older adults: a latent class analysis of an ageing Irish cohort. Public Health Nutr. 2014;17(12):2674–2686. doi:10.1017/S1368980014000111

44. Hearty AP, Gibney MJ. Analysis of meal patterns with the use of supervised data mining techniques--artificial neural networks and decision trees. Am J Clin Nutr. 2008;88(6):1632–1642. doi:10.3945/ajcn.2008.26619

45. Iqbal K, Buijsse B, Wirth J, Schulze MB, Floegel A, Boeing H. Gaussian graphical models identify networks of dietary intake in a German adult population. The Journal of Nutrition. 2016;146(3):646–652. doi:10.3945/JN.115.221135

46. Schwedhelm C, Knüppel S, Schwingshackl L, Boeing H, Iqbal K. Meal and habitual dietary networks identified through Semiparametric Gaussian Copula Graphical Models in a German adult population. PLoS One. 2018;13(8):e0202936. doi:10.1371/journal.pone.0202936

47. Solans M, Coenders G, Marcos-Gragera R, et al. Compositional analysis of dietary patterns. Stat Methods Med Res. 2019;28(9):2834–2847. doi:10.1177/0962280218790110

48. Dalmartello M, Decarli A, Ferraroni M, et al. Dietary patterns and oral and pharyngeal cancer using latent class analysis. Int J Cancer. 2020;147(3):719–727. doi:10.1002/ijc.32769

49. Hoang T, Lee J, Kim J. Network analysis of demographics, dietary intake, and comorbidity interactions. Nutrients. 2021;13(10):3563. doi:10.3390/NU13103563/S1

50. Hose AJ, Pagani G, Karvonen AM, et al. Excessive unbalanced meat consumption in the first year of life increases asthma risk in the PASTURE and LUKAS2 birth cohorts. Front Immunol. 2021;12:651709. doi:10.3389/fimmu.2021.651709

51. Madeira T, Severo M, Oliveira A, Gorjão Clara J, Lopes C. The association between dietary patterns and nutritional status in community-dwelling older adults-the PEN-3S study. Eur J Clin Nutr. 2021;75(3):521–530. doi:10.1038/s41430-020-00745-w

52. Samieri C, Sonawane AR, Lefèvre-Arbogast S, Helmer C, Grodstein F, Glass K. Using network science tools to identify novel diet patterns in prodromal dementia. Neurology. 2020;94(19):e2014–e2025. doi:10.1212/WNL.0000000000009399

53. Shang X, Li Y, Xu H, et al. Leading dietary determinants identified using machine learning techniques and a healthy diet score for changes in cardiometabolic risk factors in children: a longitudinal analysis. Nutr J. 2020;19(1):105. doi:10.1186/s12937-020-00611-2

54. Wright DM, McKenna G, Nugent A, Winning L, Linden GJ, Woodside JV. Association between diet and periodontitis: a cross-sectional study of 10,000 NHANES participants. Am J Clin Nutr. 2020;112(6):1485–1491. doi:10.1093/ajcn/nqaa266

55. Xia Y, Zhao Z, Zhang S, et al. Complex dietary topologies in non-alcoholic fatty liver disease: A network science analysis. Front Nutr. 2020;7:579086. doi:10.3389/fnut.2020.579086

56. Zhao Y, Naumova EN, Bobb JF, Claus Henn B, Singh GM. Joint associations of multiple dietary components with cardiovascular disease risk: A machine-learning approach. Am J Epidemiol. 2021;190(7):1353–1365. doi:10.1093/aje/kwab004

57. de Almeida Alves M, Molina MDCB, da Fonseca M de JM, Lotufo PA, Benseñor IM, Marchioni DML. Different statistical methods identify similar population-specific dietary patterns: an analysis of Longitudinal Study of Adult Health (ELSA-Brasil). Br J Nutr. 2022;128(11):2249–2257. doi:10.1017/S0007114522000253

58. Farmer N, Lee LJ, Powell-Wiley TM, Wallen GR. Cooking frequency and perception of diet among US adults are associated with US healthy and healthy Mediterranean-style dietary related classes: A latent class profile analysis. Nutrients. 2020;12(11):3268. doi:10.3390/nu12113268

59. Schwedhelm C, Lipsky LM, Shearrer GE, et al. Using food network analysis to understand meal patterns in pregnant women with high and low diet quality. International Journal of Behavioral Nutrition and Physical Activity. 2021;18(1):101. doi:10.1186/s12966-021-01172-1

60. Panaretos D, Koloverou E, Dimopoulos AC, et al. A comparison of statistical and machine-learning techniques in evaluating the association between dietary patterns and 10-year cardiometabolic risk (2002-2012): the ATTICA study. The British Journal of Nutrition. 2018;120(3):326–334. doi:10.1017/S0007114518001150

61. Hanson KL, Connor LM. Food insecurity and dietary quality in US adults and children: A systematic review. Am J Clin Nutr. 2014;100(2):684–692. doi:10.3945/ajcn.114.084525

62. Doan N, Olstad DL, Vanderlee L, Hammond D, Wallace M, Kirkpatrick SI. Investigating the intersections of racial identity and perceived income adequacy in relation to dietary quality among adults in Canada. The Journal of Nutrition. 2022;13(152):67S–75S. doi:10.1093/JN/NXAC076

63. Hiza HAB, Casavale KO, Guenther PM, Davis CA. Diet quality of Americans differs by age, sex, race/ethnicity, income, and education level. Journal of the Academy of Nutrition and Dietetics. 2013;113(2):297–306. doi:10.1016/J.JAND.2012.08.011

64. Olstad DL, Nejatinamini S, Victorino C, Kirkpatrick SI, Minaker LM, McLaren L. Trends in socioeconomic inequities in diet quality between 2004 and 2015 among a nationally representative sample of children in Canada. The Journal of Nutrition. 2021;151(12):3781–3794. doi:10.1093/JN/NXAB297

65. Brassard D, Munene LAE, Pierre SS, et al. Evaluation of the Healthy Eating Food Index (HEFI)-2019 measuring adherence to Canada’s Food Guide 2019 recommendations on healthy food choices. *Applied Physiology*, Nutrition, and Metabolism. 2022;47(5). doi:10.1139/APNM-2021-0416

66. Wang A, Ramaswamy VV, Russakovsky O. Towards intersectionality in machine learning: Including more identities, handling underrepresentation, and performing evaluation. In: Proceedings of the 2022 ACM Conference on Fairness, Accountability, and Transparency. FAccT ‘22. Association for Computing Machinery; 2022:336–349. doi: 10.1145/3531146.3533101

67. Robinson WR, Renson A, Naimi AI. Teaching yourself about structural racism will improve your machine learning. Biostatistics. 2020;21(2):339–344. doi:10.1093/biostatistics/kxz040

68. Rojas JC, Fahrenbach J, Makhni S, et al. Framework for integrating equity into machine learning models: A case study. Chest. 2022;161(6):1621–1627. doi:10.1016/j.chest.2022.02.001

69. Rajkomar A, Hardt M, Howell MD, Corrado G, Chin MH. Ensuring fairness in machine learning to advance health equity. Annals of Internal Medicine. 2018;169(12):866–872. doi:10.7326/M18-1990

70. Lachat C, Hawwash D, Ocké MC, et al. Strengthening the Reporting of Observational Studies in Epidemiology-Nutritional Epidemiology (STROBE-nut): An extension of the STROBE statement. PLOS Medicine. 2016;13(6). doi:10.1371/JOURNAL.PMED.1002036

71. Liu X, Cruz Rivera S, Moher D, et al. Reporting guidelines for clinical trial reports for interventions involving artificial intelligence: the CONSORT-AI extension. Nature Medicine. 2020;26(9):1364–1374. doi:10.1038/s41591-020-1034-x

72. Spicker D, Nazemi A, Hutchinson J, et al. Challenges for predictive modeling with neural network techniques using error-prone dietary intake data. Published online November 15, 2023. doi:10.48550/arXiv.2311.09338

73. Lampignano L, Tatoli R, Donghia R, et al. Nutritional patterns as machine learning predictors of liver health in a population of elderly subjects. Nutr Metab Cardiovasc Dis. 2023;33(11):2233–2241. doi:10.1016/j.numecd.2023.07.009

74. Slurink IA, Corpeleijn E, Bakker SJ, et al. Dairy consumption and incident prediabetes: prospective associations and network models in the large population-based Lifelines Study. Am J Clin Nutr. 2023;118(6):1077–1090. doi:10.1016/j.ajcnut.2023.10.002

75. Kirk D, Catal C, Tekinerdogan B. Precision nutrition: A systematic literature review. Computers in Biology and Medicine. 2021;133:104365. doi:10.1016/j.compbiomed.2021.104365

76. Breiman L. Random Forests. Machine Learning. 2001;45(1):5-32. doi:10.1023/A:1010933404324

77. Song Y yan, Lu Y. Decision tree methods: applications for classification and prediction. Shanghai Arch Psychiatry. 2015;27(2):130–135. doi:10.11919/j.issn.1002-0829.215044

78. Grossi E, Buscema M. Introduction to artificial neural networks. European Journal of Gastroenterology & Hepatology. 2007;19(12):1046. doi:10.1097/MEG.0b013e3282f198a0

79. Bianchi D, Calogero R, Tirozzi B. Kohonen neural networks and genetic classification. Mathematical and Computer Modelling. 2007;45(1):34–60. doi:10.1016/j.mcm.2006.04.004

80. Drton M, Maathuis MH. Structure learning in graphical modeling. 101146/annurev-statistics-060116-053803. 2017;4:365–393. doi:10.1146/ANNUREV-STATISTICS-060116-053803

81. Altenbuchinger M, Weihs A, Quackenbush J, Grabe HJ, Zacharias HU. Gaussian and Mixed Graphical Models as (multi-)omics data analysis tools. Biochimica et Biophysica Acta (BBA) – Gene Regulatory Mechanisms. 2020;1863(6):194418. doi:10.1016/J.BBAGRM.2019.194418

82. Koller D, Friedman N. Probabilistic Graphical Models: Principles and Techniques. MIT Press; 2009.

83. Liu H, Han F, Yuan M, Lafferty J, Wasserman L. High-dimensional semiparametric Gaussian copula graphical models. The Annals of Statistics. 2012;40(4):2293–2326.

84. Freund Y, Schapire RE. A decision-theoretic generalization of on-line learning and an application to boosting. Journal of Computer and System Sciences. 1997;55(1):119–139. doi:10.1006/jcss.1997.1504

85. Bobb JF, Valeri L, Claus Henn B, et al. Bayesian kernel machine regression for estimating the health effects of multi-pollutant mixtures. Biostatistics. 2015;16(3):493–508. doi:10.1093/biostatistics/kxu058

86. Aitchison J. The statistical analysis of compositional data. Journal of the Royal Statistical Society Series B (Methodological). 1982;44(2):139–177.

87. Rodrigues LA, Daunis-i-Estadella P, Figueras G, Henestrosa S. Flying in compositional morphospaces: evolution of limb proportions in flying vertibrates. In: Compositional Data Analysis: Theory and Applications.; 2011:235–254. doi:10.1002/9781119976462.ch17

88. Zhang CH, Huang J. The sparsity and bias of the Lasso selection in high-dimensional linear regression. The Annals of Statistics. 2008;36(4):1567–1594.

89. Oliveira A, Lopes C, Torres D, Ramos E, Severo M. Application of a latent transition model to estimate the usual prevalence of dietary patterns. Nutrients. 2021;13(1):133. doi:10.3390/nu13010133

90. Fahey MT, Thane CW, Bramwell GD, Coward WA. Conditional Gaussian mixture modelling for dietary pattern analysis. Journal of the Royal Statistical Society Series A (Statistics in Society). 2007;170(1):149–166.

91. Cover T, Thomas J. Elements of Information Theory. Wiley; 2005. Accessed August 23, 2023. https://books-scholarsportal-info.proxy.lib.uwaterloo.ca/en/read?id=/ebooks/ebooks2/pda/2011-12-01/1/13568.9780471748816#page=1

92. Gorst-Rasmussen A, Dahm C, Dethlefsen C, Scheike T, Overvad K. Exploring dietary patterns by using the treelet transform. Am J Epidemiol. 2011;173:1097–1104. doi:10.1093/aje/kwr060

93. Lee AB, Nadler B, Wasserman L. Treelets—An adaptive multi-scale basis for sparse unordered data. The Annals of Applied Statistics. 2008;2(2):435–471. doi:10.1214/07-AOAS137

