## Supplemental File 1 for "Advances in methods for characterizing dietary patterns: A scoping review"

### Supplemental File 1: Search strategies for MEDLINE, CINAHL, and SCOPUS

Due to the emerging nature of this field, an updated search of the literature was conducted in March 2022. The search for relevant articles was applied in the same three academic databases: MEDLINE, CINAHL, and Scopus. No date limits were applied to the searches and the same search was applied with minor changes, such as the removal of body weight and reduced rank regression.

**MEDLINE 2019 search**

((systems modelling [tiab] OR systems science [tiab] OR systems thinking [tiab] OR systems approach*[tiab] OR systems theory*[tiab] OR systems analysis [tiab] OR agent-based model*[tiab] OR system dynamics model*[tiab] OR systems-based [tiab]) OR (LASSO [tiab] OR "least absolute shrinkage and selection operator" [tiab]) OR (reduced rank regression [tiab] OR RRR [tiab]) OR (machine learning [tiab] OR neural networks [tiab] OR decision tree analys* [tiab] OR deep learning [tiab] OR deep neural networks [tiab] OR network science [tiab] OR network analys* [tiab] OR topological data analys* [tiab] OR network-based [tiab]) OR (copula* [tiab] OR copular function* [tiab]) OR ("data driven" [tiab] OR posteriori [tiab] OR priori [tiab] OR "hypothesis driven" [tiab]))

AND

(eating pattern*[tiab] OR diet pattern*[tiab] OR dietary pattern*[tiab] OR food pattern*[tiab] OR food habit*[tiab] OR diet quality score*[tiab] OR diet ind*[tiab] OR dietary ind*[tiab] OR dietary guide*[tiab] OR dietary recommend* [tiab] OR diet scor* [tiab] OR "healthy eating index" [tiab] OR dietary intake*[tiab] OR diet intake*[tiab] OR obes*[tiab] OR body weight [tiab])

Filter: Humans; No date limiter.

**MEDLINE 2022 search with no body weight terms**
((systems modelling [tiab] OR systems science [tiab] OR systems thinking [tiab] OR systems approach*[tiab] OR systems theory*[tiab] OR systems analysis [tiab] OR agent-based model*[tiab] OR system dynamics model*[tiab] OR systems-based [tiab]) OR (LASSO [tiab] OR "least absolute shrinkage and selection operator" [tiab]) OR (reduced rank regression [tiab] OR RRR [tiab]) OR (machine learning [tiab] OR neural networks [tiab] OR decision tree analys* [tiab] OR deep learning [tiab] OR deep neural networks [tiab] OR network science [tiab] OR network analys* [tiab] OR topological data analys* [tiab] OR network-based [tiab]) OR (copula* [tiab] OR copular function* [tiab]) OR ("data driven" [tiab] OR posteriori [tiab] OR priori [tiab] OR "hypothesis driven" [tiab]))

AND

(feeding behavior[mesh:noexp] OR eating pattern*[tiab] OR diet pattern*[tiab] OR dietary pattern*[tiab] OR food pattern*[tiab] OR food habit*[tiab] OR diet quality score*[tiab] OR diet ind*[tiab] OR dietary ind*[tiab] OR dietary guide*[tiab] OR dietary recommend* [tiab] OR diet scor* [tiab] OR "healthy eating index" [tiab] OR dietary intake*[tiab] OR diet intake*[tiab])

Filter: Humans; Date limiter: AND ("2019/10/25"[Date - Entry] : "3000"[Date - Entry])

**MEDLINE 2022 search with subject heading added**
(((systems modelling [tiab] OR systems science [tiab] OR systems thinking [tiab] OR systems approach*[tiab] OR systems theory*[tiab] OR systems analysis [tiab] OR agent-based model*[tiab] OR system dynamics model*[tiab] OR systems-based [tiab]) OR (LASSO [tiab] OR "least absolute shrinkage and selection operator" [tiab]) OR (reduced rank regression [tiab] OR RRR [tiab]) OR (machine learning [tiab] OR neural networks [tiab] OR decision tree analys* [tiab] OR deep learning [tiab] OR deep neural networks [tiab] OR network science [tiab] OR network analys* [tiab] OR topological data analys* [tiab] OR network-based [tiab]) OR (copula* [tiab] OR copular function* [tiab]) OR ("data driven" [tiab] OR posteriori [tiab] OR priori [tiab] OR "hypothesis driven" [tiab]))

AND

(feeding behavior[mesh:noexp] OR eating pattern*[tiab] OR diet pattern*[tiab] OR dietary pattern*[tiab] OR food pattern*[tiab] OR food habit*[tiab] OR diet quality score*[tiab] OR diet ind*[tiab] OR dietary ind*[tiab] OR dietary guide*[tiab] OR dietary recommend* [tiab] OR diet scor* [tiab] OR "healthy eating index" [tiab] OR dietary intake*[tiab] OR diet intake*[tiab] OR obes*[tiab] OR body weight [tiab]))

NOT

(((systems modelling [tiab] OR systems science [tiab] OR systems thinking [tiab] OR systems approach*[tiab] OR systems theory*[tiab] OR systems analysis [tiab] OR agent-based model*[tiab] OR system dynamics model*[tiab] OR systems-based [tiab]) OR (LASSO [tiab] OR "least absolute shrinkage and selection operator" [tiab]) OR (reduced rank regression [tiab] OR RRR [tiab]) OR (machine learning [tiab] OR neural networks [tiab] OR decision tree analys* [tiab] OR deep learning [tiab] OR deep neural networks [tiab] OR network science [tiab] OR network analys* [tiab] OR topological data analys* [tiab] OR network-based [tiab]) OR (copula* [tiab] OR copular function* [tiab]) OR ("data driven" [tiab] OR posteriori [tiab] OR priori [tiab] OR "hypothesis driven" [tiab]))

AND

(eating pattern*[tiab] OR diet pattern*[tiab] OR dietary pattern*[tiab] OR food pattern*[tiab] OR food habit*[tiab] OR diet quality score*[tiab] OR diet ind*[tiab] OR dietary ind*[tiab] OR dietary guide*[tiab] OR dietary recommend* [tiab] OR diet scor* [tiab] OR "healthy eating index" [tiab] OR dietary intake*[tiab] OR diet intake*[tiab] OR obes*[tiab] OR body weight [tiab])

Filter: Humans; No date limiter.

**CINAHL 2019 search**

TI ( eating pattern* OR diet pattern* OR dietary pattern* OR food pattern* OR food habit* OR diet quality score* OR diet ind* OR dietary ind* OR dietary guide* OR dietary recommend* OR diet scor* OR “healthy eating index” OR dietary intake* OR diet intake* OR obes* OR body weight) OR AB ( eating pattern* OR diet pattern* OR dietary pattern* OR food pattern* OR food habit* OR diet quality score* OR diet ind* OR dietary ind* OR dietary guide* OR dietary recommend* OR diet scor* OR “healthy eating index” OR dietary intake* OR diet intake* OR obes* OR body weight)

AND

TI ( systems modelling OR systems science OR systems thinking OR systems approach* OR systems theory* OR systems analysis OR agent-based model* OR system dynamics model* OR systems-based ) OR AB ( systems modelling OR systems science OR systems thinking OR systems approach* OR systems theory* OR systems analysis OR agent-based model* OR system dynamics model* OR systems-based ) ) OR ( TI ( LASSO OR “least absolute shrinkage and selection operator” ) OR AB ( LASSO OR “least absolute shrinkage and selection operator” ) ) OR ( TI ( reduced rank regression OR RRR ) OR AB ( reduced rank regression OR RRR ) ) OR ( TI ( machine learning OR neural networks OR decision tree analys* OR deep learning OR deep neural networks OR network science OR network analys* OR topological data analys* OR network-based ) OR AB ( machine learning OR neural networks OR decision tree analys* OR deep learning OR deep neural networks OR network science OR network analys* OR topological data analys* OR network-based ) ) OR ( TI ( copula* OR copular function* ) OR AB ( copula* OR copular function* ) ) OR ( TI ( “data driven” OR posteriori OR priori OR “hypothesis driven” ) OR AB ( “data driven” OR posteriori OR priori OR “hypothesis driven” )

Filter: Human; No date limiter.

**CINAHL 2022 search with no body weight terms**

TI ( eating pattern* OR diet pattern* OR dietary pattern* OR food pattern* OR food habit* OR diet quality score* OR diet ind* OR dietary ind* OR dietary guide* OR dietary recommend* OR diet scor* OR “healthy eating index” OR dietary intake* OR diet intake*) OR AB ( eating pattern* OR diet pattern* OR dietary pattern* OR food pattern* OR food habit* OR diet quality score* OR diet ind* OR dietary ind* OR dietary guide* OR dietary recommend* OR diet scor* OR “healthy eating index” OR dietary intake* OR diet intake*)

AND

TI ( systems modelling OR systems science OR systems thinking OR systems approach* OR systems theory* OR systems analysis OR agent-based model* OR system dynamics model* OR systems-based ) OR AB ( systems modelling OR systems science OR systems thinking OR systems approach* OR systems theory* OR systems analysis OR agent-based model* OR system dynamics model* OR systems-based ) ) OR ( TI ( LASSO OR “least absolute shrinkage and selection operator” ) OR AB ( LASSO OR “least absolute shrinkage and selection operator” ) ) OR ( TI ( reduced rank regression OR RRR ) OR AB ( reduced rank regression OR RRR ) ) OR ( TI ( machine learning OR neural networks OR decision tree analys* OR deep learning OR deep neural networks OR network science OR network analys* OR topological data analys* OR network-based ) OR AB ( machine learning OR neural networks OR decision tree analys* OR deep learning OR deep neural networks OR network science OR network analys* OR topological data analys* OR network-based ) ) OR ( TI ( copula* OR copular function* ) OR AB ( copula* OR copular function* ) ) OR ( TI ( “data driven” OR posteriori OR priori OR “hypothesis driven” ) OR AB ( “data driven” OR posteriori OR priori OR “hypothesis driven” )

Filter: Human; Date limiter: AND ZD (201910* OR 202* OR "in process")

**CINAHL 2022 search with subject heading added**

TI ( eating pattern* OR diet pattern* OR dietary pattern* OR food pattern* OR food habit* OR diet quality score* OR diet ind* OR dietary ind* OR dietary guide* OR dietary recommend* OR diet scor* OR “healthy eating index” OR dietary intake* OR diet intake* OR obes* OR body weight) OR AB ( eating pattern* OR diet pattern* OR dietary pattern* OR food pattern* OR food habit* OR diet quality score* OR diet ind* OR dietary ind* OR dietary guide* OR dietary recommend* OR diet scor* OR “healthy eating index” OR dietary intake* OR diet intake* OR obes* OR body weight) OR MH (food habits)

NOT

TI ( eating pattern* OR diet pattern* OR dietary pattern* OR food pattern* OR food habit* OR diet quality score* OR diet ind* OR dietary ind* OR dietary guide* OR dietary recommend* OR diet scor* OR “healthy eating index” OR dietary intake* OR diet intake* OR obes* OR body weight) OR AB ( eating pattern* OR diet pattern* OR dietary pattern* OR food pattern* OR food habit* OR diet quality score* OR diet ind* OR dietary ind* OR dietary guide* OR dietary recommend* OR diet scor* OR “healthy eating index” OR dietary intake* OR diet intake* OR obes* OR body weight)

AND

TI ( systems modelling OR systems science OR systems thinking OR systems approach* OR systems theory* OR systems analysis OR agent-based model* OR system dynamics model* OR systems-based ) OR AB ( systems modelling OR systems science OR systems thinking OR systems approach* OR systems theory* OR systems analysis OR agent-based model* OR system dynamics model* OR systems-based ) ) OR ( TI ( LASSO OR “least absolute shrinkage and selection operator” ) OR AB ( LASSO OR “least absolute shrinkage and selection operator” ) ) OR ( TI ( reduced rank regression OR RRR ) OR AB ( reduced rank regression OR RRR ) ) OR ( TI ( machine learning OR neural networks OR decision tree analys* OR deep learning OR deep neural networks OR network science OR network analys* OR topological data analys* OR network-based ) OR AB ( machine learning OR neural networks OR decision tree analys* OR deep learning OR deep neural networks OR network science OR network analys* OR topological data analys* OR network-based ) ) OR ( TI ( copula* OR copular function* ) OR AB ( copula* OR copular function* ) ) OR ( TI ( “data driven” OR posteriori OR priori OR “hypothesis driven” ) OR AB ( “data driven” OR posteriori OR priori OR “hypothesis driven” )

Filter: Human; No date limiter.

**Scopus 2019 search**
TITLE-ABS-KEY("eating pattern" OR "diet pattern" OR "dietary pattern" OR "food pattern" OR "food habit" OR "diet quality score" OR "diet index" OR "dietary index" OR "dietary guide" OR "dietary recommend" OR "diet score" OR “healthy eating index” OR "dietary intake" OR "diet intake" OR obes OR “body weight”)

AND

TITLE-ABS-KEY("systems modelling" OR "systems science" OR "systems thinking" OR "systems approach" OR "systems theory" OR "systems analysis" OR "agent-based model" OR "system dynamics model" OR "systems-based" OR LASSO OR “least absolute shrinkage and selection operator” OR "reduced rank regression" OR RRR OR "machine learning" OR "neural networks" OR "decision tree analysis" OR "deep learning" OR "deep neural networks" OR "network science" OR "network analysis" OR "topological data analysis" OR "network-based" OR "copula" OR "copular function" OR “data driven” OR "posteriori" OR "priori" OR “hypothesis driven”)

No date limiter.

**Scopus 2022 search with no body weight terms**

TITLE-ABS-KEY("eating pattern" OR "diet pattern" OR "dietary pattern" OR "food pattern" OR "food habit" OR "diet quality score" OR "diet index"  OR "dietary index" OR "dietary guide" OR "dietary recommend"  OR "diet score"  OR “healthy eating index”  OR "dietary intake" OR "diet intake")

AND

TITLE-ABS-KEY("systems modelling" OR "systems science" OR "systems thinking" OR "systems approach" OR "systems theory" OR "systems analysis" OR "agent-based model" OR "system dynamics model" OR "systems-based" OR LASSO OR “least absolute shrinkage and selection operator” OR "reduced rank regression" OR RRR OR "machine learning" OR "neural networks"  OR "decision tree analysis" OR "deep learning" OR "deep neural networks" OR "network science" OR "network analysis" OR "topological data analysis" OR "network-based" OR "copula" OR "copular function" OR “data driven” OR "posteriori" OR "priori" OR “hypothesis driven”)

Date limiter: 2019 to present
