## Supplemental File 2 for "Advances in methods for characterizing dietary patterns: A scoping review"

### Supplemental File 2: Key extraction fields for review of novel analytic methods to characterize dietary patterns

| Field | Description |
|---|---|
| Study identification | Authors |
| | Title |
| | Journal |
| | Year published |
| | Funding source for study |
| Study population | Sample size |
| | Sample characteristics |
| Methods | Research objectives/questions |
| | Study design |
| | Study name |
| | Measurement of diet |
| | Dietary components analyzed |
| | Novel methods of analysis |
| | Consideration of equity |
| Results | Findings re: diet or diet-related outcomes |
| | Author description of method utility and implications |
| Critical review | Author stated limitations |
| | Reviewer observed limitations |
